# A fast and accurate method for estimating the proportion of asymptomatic infections from serosurveys: Application to SARS-CoV-2 and beyond

**DOI:** 10.1101/2024.09.11.24313462

**Authors:** Akshay Tiwari, Shreya Chowdhury, Ananthu James, Budhaditya Chatterjee, Narendra M. Dixit

## Abstract

Several pathogens, including SARS-CoV-2, can cause asymptomatic infections which remain undetected but transmit disease. Estimating the proportion of asymptomatic infections, *ψ*, is therefore important for epidemic control. Accurate estimation of *ψ* has been challenging. Population-based surveys misclassify asymptomatic individuals as symptomatic when they experience overlapping symptoms from unrelated conditions. Here, we developed a rigorous mathematical formalism that accounts for symptom overlap together with infection test sensitivity and specificity and yields accurate estimates of *ψ*. We validated the formalism against synthetic datasets. We then applied it to 50 COVID-19 serosurveys, spanning ∼0.8 million individuals across 29 nations. *ψ* was significantly higher (median ∼60%) than previously reported (∼40%) (P=8×10^-11^). Symptom overlap explained ∼89% of this difference. Furthermore, *ψ* became more consistent within nations, helping better devise intervention policies. We applied our formalism also to Zika virus outbreaks, demonstrating its broad applicability. Finally, we developed a web-application for its easy implementation (URL: https://therapeuticengglab.github.io/ShinyPsi/).

## INTRODUCTION

Asymptomatic infections, a key feature of several globally important pathogens^1-7^, pose a major challenge to epidemic control measures. They remain undetected and silently fuel disease spread.^1^ Asymptomatic SARS-CoV-2 infections, for instance, contributed enormously to the COVID-19 pandemic: depending on the prevalent conditions, between ∼3% and ∼69% of all transmission events have been attributed to them.^8,9^ Knowledge of their proportion, *ψ*, namely, the fraction of infections that remains asymptomatic, is thus important for understanding disease epidemiology, forecasting future outbreaks, and designing and assessing public health interventions.^8,10-15^ Accurate estimation of *ψ*, however, has been a challenge.

Current estimation methods, employed from the early days of the pandemic^16,17^, rely on population-based surveys that contain two parts: 1) a nucleic acid or an antibody test to detect current or past infections, respectively, and 2) a questionnaire to assess the symptoms experienced. Individuals who test positive for the infection but declare no symptoms are deemed asymptomatically infected. *ψ* is thus estimated as the fraction of test-positive individuals that reports no symptoms. This method has an important limitation: In the context of SARS-CoV-2, for instance, symptoms such as cough and fever, which are part of nearly all COVID-19 surveys, can be triggered not only by SARS-CoV-2 infection but also by a host of other conditions including infections by influenza virus or other circulating coronaviruses. It is possible, therefore, that some individuals who report symptoms in the surveys may have experienced them due to the other conditions. Such individuals should be classified as asymptomatic for SARS-CoV-2 but get misclassified as symptomatic, resulting in a systematic underestimation of *ψ*.

The possibility of this misclassification is evident in the data gathered by the surveys: The surveys identify individuals who test negative for SARS-CoV-2 but report symptoms. For instance, a survey from The Netherlands reported that ∼62% of the individuals who tested negative for SARS-CoV-2 displayed symptoms.^18^ The number was as high as 80% in surveys in the US.^19,20^ These individuals, barring false negatives, must have had their symptoms arise from causes other than SARS-CoV-2 infection. The high proportion of such individuals in these surveys implies that at least some of the test-positive, symptomatic individuals may have had their symptoms arise from non-COVID-19 conditions. This phenomenon is evident with other pathogens too.^7^ Accounting for this phenomenon is important to obtain accurate estimates of *ψ*.

The challenge arises from the two-part survey methodology, in which misclassification from symptom overlap with other conditions gets entangled with misidentification of infection status due to the imperfect sensitivity and specificity of the infection test. Thus, the test-negative, symptomatic individuals discussed above may not all have been uninfected; some who had the infection may have been classified as test-negative because the antigen (or antibody) levels in them were below assay detection limits. Indeed, the symptoms they experienced may well have arisen from SARS-CoV-2 infection. Thus, accounting for symptom overlap must also simultaneously contend with the sensitivity and specificity of the infection test, which is challenging.

Previous studies have attempted to address this challenge but met with limited success. In the late 1970s, a general formalism to account for antibody (or nucleic acid) test sensitivity and specificity was developed, yielding the so called Rogan-Gladen estimator of disease prevalence.^21^ The formalism has been applied to estimate SARS-CoV-2 prevalence during the pandemic.^18^ It was later extended to estimate *ψ*, but without accounting for symptom overlap.^22^ Contrastingly, a study estimated *ψ* by accounting for symptom overlap but not test sensitivity and specificity.^23^

In an important advance made in the late 2010s, just before the pandemic, a study developed a Bayesian inference framework to estimate the proportion of symptomatic infections (i.e., 1 − *ψ*) by accounting for test sensitivity and specificity as well as symptom overlap in the context of Zika virus infection.^7^ The method could, in principle, be used to analyze data from population-based surveys and estimate *ψ*. It, however, requires complex Markov-chain-Monte-Carlo (MCMC) simulations, which make its implementation involved and computationally expensive. Furthermore, the complex nature of the method makes interpretation of the results difficult. Indeed, the method has not been applied to any SARS-CoV-2 dataset, despite the hundreds of serosurveys reported during the pandemic.^24^ Estimates of *ψ* for SARS-CoV-2 thus remain uncertain. A method for estimating *ψ* that is efficient, transparent, and accurate, for SARS-CoV-2 as well as other pathogens, is thus needed.

Here, we developed a rigorous mathematical formalism that not only accounts simultaneously for test sensitivity and specificity and symptom overlap, allowing accurate estimation of *ψ* from standard two-part population-based surveys, but also yields an analytical, closed-form expression for *ψ*, enabling its ready application and interpretation. We validated the formalism against synthetic datasets and then applied it to data from 50 COVID-19 and 3 Zika virus serosurveys. Finally, we developed a freely available web application for easy implementation of our formalism that uses standard data collected in serosurveys as inputs and yields estimates of *ψ* in seconds.

## RESULTS

### Formalism to estimate the proportion of asymptomatic infections

We considered the general scenario where the symptoms triggered by the pathogen of interest could also be triggered by other pathogens/conditions prevalent in the population. We assumed that data associated with the pathogen of interest is gathered following the two-part survey methodology described above. We developed a formalism to estimate *ψ*, the proportion of infections asymptomatic for the pathogen of interest (Methods). The formalism accounted for the sensitivity and specificity of the infection test as well as the effect of symptom overlap with other conditions. It yielded the following closed-form, analytical formula for *ψ*:

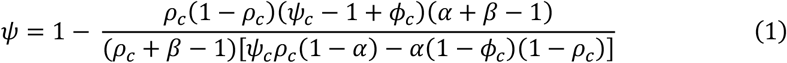

Here, *α* and *β* are the test sensitivity and specificity, respectively; *ψ*_*c*_ is the crude (or reported) proportion of asymptomatic individuals among test-positive individuals; *ρ*_*c*_ is the crude seroprevalence, the fraction of test-positive individuals among the sampled individuals; and *ϕ*_*c*_ is the crude proportion of symptomatic individuals among test-negative individuals. Thus, given the set of quantities *Q =* }*α, β, ρ*_*c*_, *ϕ*_*c*_, *ψ*_*c*_}, all of which are typically reported in surveys, *ψ* can be calculated.

To understand the dependence of *ψ* on each of the quantities in *Q*, we solved equation (1) by varying values of the latter quantities over ranges representative of SARS-CoV-2 serosurveys (see below). We thus chose three combinations of *α* and *β* values, representing antibody tests with low, moderate, and high sensitivity and specificity, respectively; three values of *ρ*_*c*_, again representing low, moderate, and high seroprevalence; and *ϕ*_*c*_ and *ψ*_*c*_ values varied over their observed ranges (Table 1). We constrained *ψ*_*c*_ to be smaller than 1 − *ϕ*_*c*_ because more asymptomatic individuals are expected in the test-negative than the test-positive subpopulation. We found that the effects of *α, β* and *ρ*_*c*_ were modest over the ranges considered compared to those of *ϕ*_*c*_ and *ψ*_*c*_ on *ψ* (Fig. 1). Increasing *ψ*_*c*_ increased *ψ*, implying, expectedly, that higher crude asymptomatic proportions were reflective of higher asymptomatic proportions overall. Importantly, increasing *ϕ*_*c*_ increased *ψ* even with constant *ψ*_*c*_. Higher *ϕ*_*c*_ implied a greater proportion of symptomatic individuals in the test-negative group, which in turn implied a greater chance of misclassification of asymptomatic individuals as symptomatic in the test-positive group. Our formalism corrected for this misclassification, resulting in higher values of *ψ*. Our formalism also helps better understand how variations in *α* and *β* with the time from infection, particularly important with nucleic acid-based tests^25,26^, may influence the accuracy of survey results (Text S1, Fig. S1). We next devised a strategy to apply the formalism to serosurvey data, accounting for the uncertainties that arise from sampling.

**Table 1.**
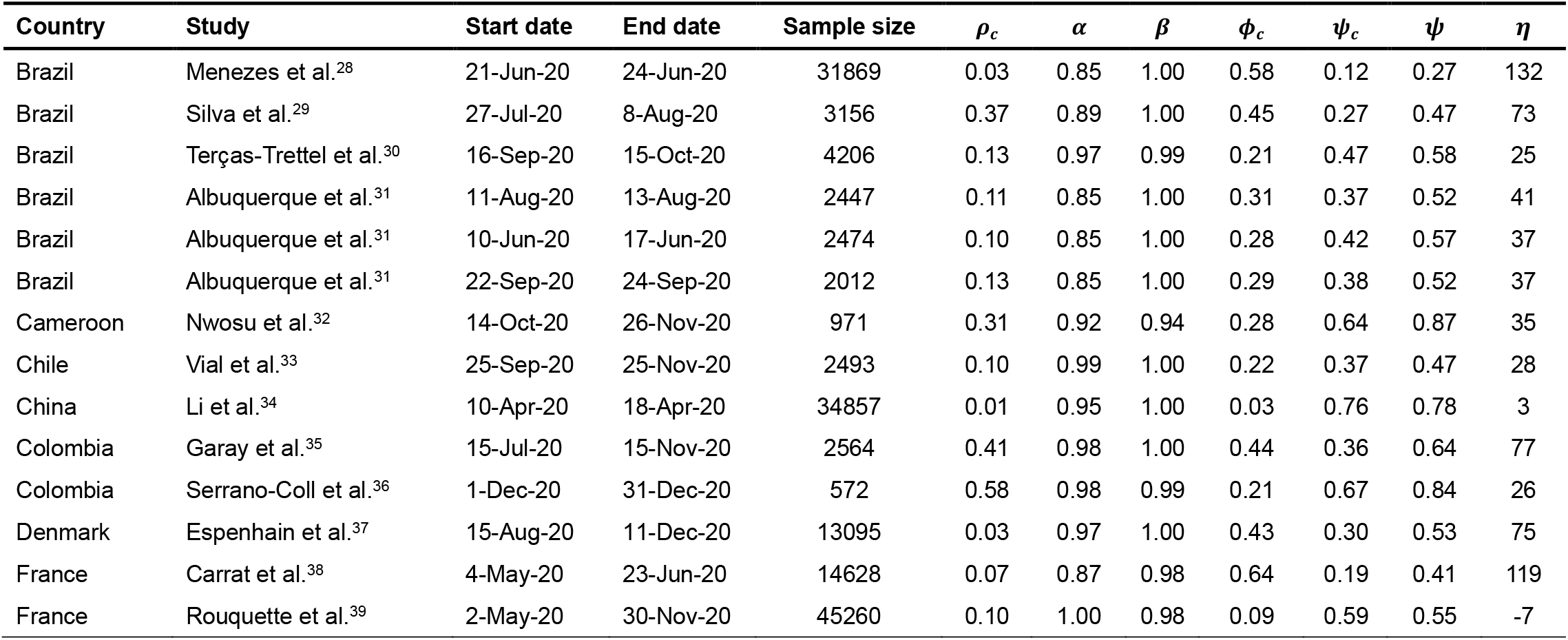
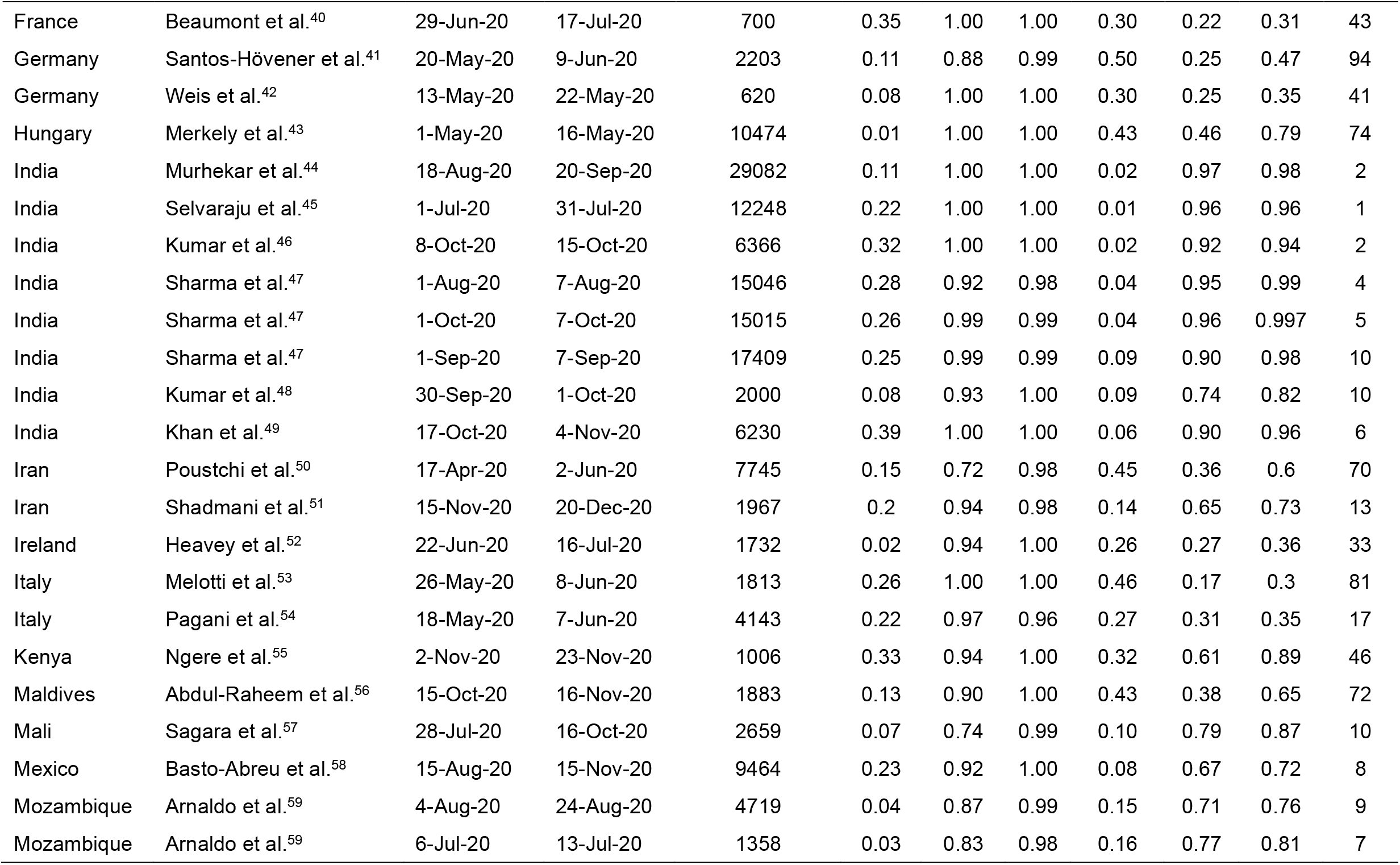
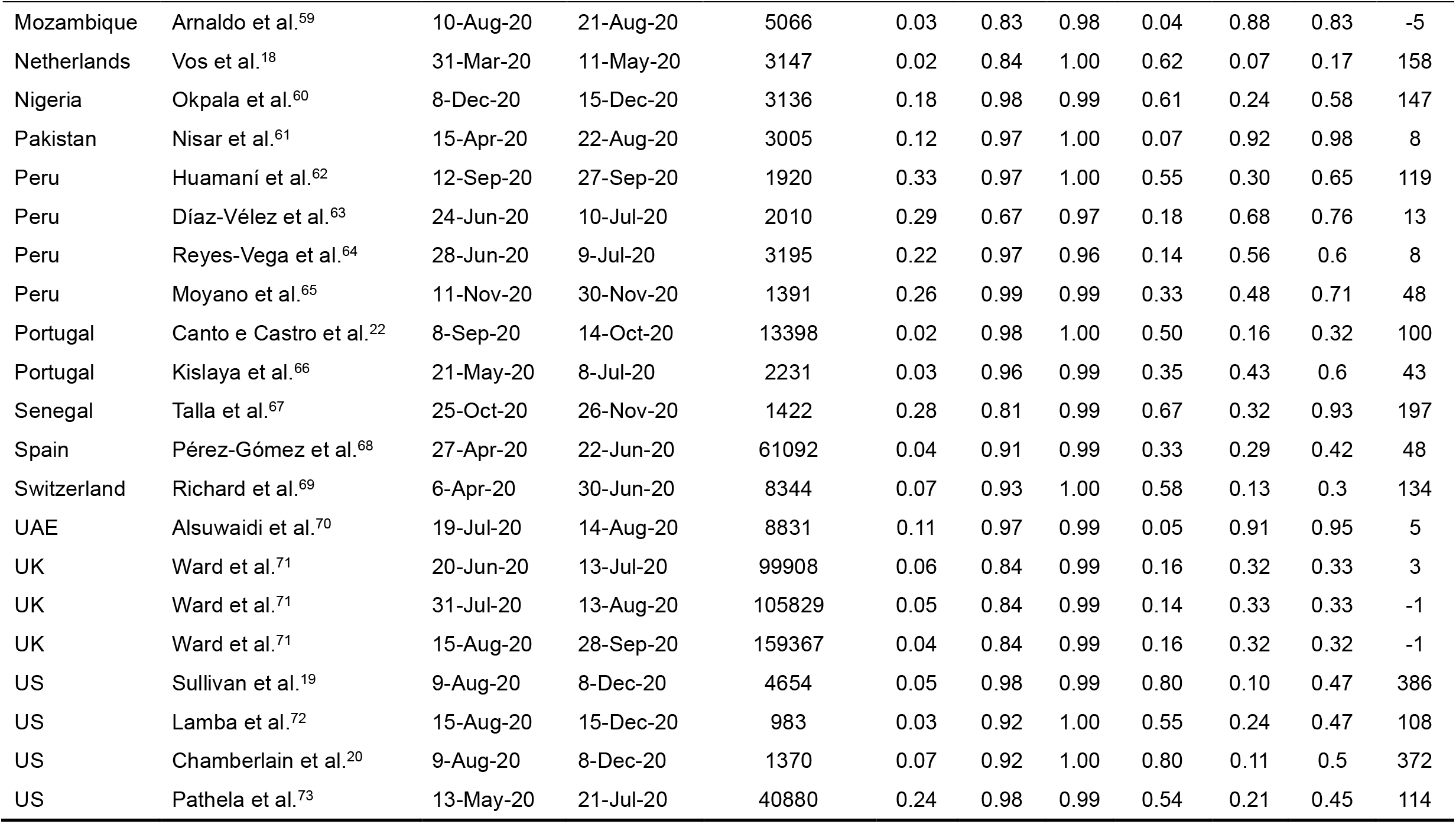
Data collated from serosurveys and our estimates of the proportion of asymptomatic infections. For each serosurvey, the country of study, the study period, sample size, reported seroprevalence (*ρ*_*c*_), test sensitivity (*α*), test specificity (*β*), proportion of symptomatic individuals among seronegative individuals (*ϕ*_*c*_), and crude (or reported) proportion of asymptomatic individuals among seropositive individuals (*ψ*_*c*_) are listed. Also listed are our estimates of the corrected proportion of asymptomatic individuals among infected individuals (*ψ*) and the extent of correction, 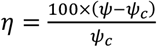. Additional details of the surveys, including their geographical scope and location, median age of participants, the symptom recall period, and test assay characteristics are in Tables S1 and S3.

| Country | Study | Start date | End date | Sample size | $\rho_c$ | $\alpha$ | $\beta$ | $\phi_c$ | $\psi_c$ | $\psi$ | $\eta$ |
| --- | --- | --- | --- | --- | --- | --- | --- | --- | --- | --- | --- |
| Brazil | Menezes et al.<sup>28</sup> | 21-Jun-20 | 24-Jun-20 | 31869 | 0.03 | 0.85 | 1.00 | 0.58 | 0.12 | 0.27 | 132 |
| Brazil | Silva et al.<sup>29</sup> | 27-Jul-20 | 8-Aug-20 | 3156 | 0.37 | 0.89 | 1.00 | 0.45 | 0.27 | 0.47 | 73 |
| Brazil | Terças-Trettel et al.<sup>30</sup> | 16-Sep-20 | 15-Oct-20 | 4206 | 0.13 | 0.97 | 0.99 | 0.21 | 0.47 | 0.58 | 25 |
| Brazil | Albuquerque et al.<sup>31</sup> | 11-Aug-20 | 13-Aug-20 | 2447 | 0.11 | 0.85 | 1.00 | 0.31 | 0.37 | 0.52 | 41 |
| Brazil | Albuquerque et al.<sup>31</sup> | 10-Jun-20 | 17-Jun-20 | 2474 | 0.10 | 0.85 | 1.00 | 0.28 | 0.42 | 0.57 | 37 |
| Brazil | Albuquerque et al.<sup>31</sup> | 22-Sep-20 | 24-Sep-20 | 2012 | 0.13 | 0.85 | 1.00 | 0.29 | 0.38 | 0.52 | 37 |
| Cameroon | Nwosu et al.<sup>32</sup> | 14-Oct-20 | 26-Nov-20 | 971 | 0.31 | 0.92 | 0.94 | 0.28 | 0.64 | 0.87 | 35 |
| Chile | Vial et al.<sup>33</sup> | 25-Sep-20 | 25-Nov-20 | 2493 | 0.10 | 0.99 | 1.00 | 0.22 | 0.37 | 0.47 | 28 |
| China | Li et al.<sup>34</sup> | 10-Apr-20 | 18-Apr-20 | 34857 | 0.01 | 0.95 | 1.00 | 0.03 | 0.76 | 0.78 | 3 |
| Colombia | Garay et al.<sup>35</sup> | 15-Jul-20 | 15-Nov-20 | 2564 | 0.41 | 0.98 | 1.00 | 0.44 | 0.36 | 0.64 | 77 |
| Colombia | Serrano-Coll et al.<sup>36</sup> | 1-Dec-20 | 31-Dec-20 | 572 | 0.58 | 0.98 | 0.99 | 0.21 | 0.67 | 0.84 | 26 |
| Denmark | Espenhain et al.<sup>37</sup> | 15-Aug-20 | 11-Dec-20 | 13095 | 0.03 | 0.97 | 1.00 | 0.43 | 0.30 | 0.53 | 75 |
| France | Carrat et al.<sup>38</sup> | 4-May-20 | 23-Jun-20 | 14628 | 0.07 | 0.87 | 0.98 | 0.64 | 0.19 | 0.41 | 119 |
| France | Rouquette et al.<sup>39</sup> | 2-May-20 | 30-Nov-20 | 45260 | 0.10 | 1.00 | 0.98 | 0.09 | 0.59 | 0.55 | -7 |
| France | Beaumont et al.<sup>40</sup> | 29-Jun-20 | 17-Jul-20 | 700 | 0.35 | 1.00 | 1.00 | 0.30 | 0.22 | 0.31 | 43 |
| Germany | Santos-Hövenner et al.<sup>41</sup> | 20-May-20 | 9-Jun-20 | 2203 | 0.11 | 0.88 | 0.99 | 0.50 | 0.25 | 0.47 | 94 |
| Germany | Weis et al.<sup>42</sup> | 13-May-20 | 22-May-20 | 620 | 0.08 | 1.00 | 1.00 | 0.30 | 0.25 | 0.35 | 41 |
| Hungary | Merkely et al.<sup>43</sup> | 1-May-20 | 16-May-20 | 10474 | 0.01 | 1.00 | 1.00 | 0.43 | 0.46 | 0.79 | 74 |
| India | Murhekar et al.<sup>44</sup> | 18-Aug-20 | 20-Sep-20 | 29082 | 0.11 | 1.00 | 1.00 | 0.02 | 0.97 | 0.98 | 2 |
| India | Selvaraju et al.<sup>45</sup> | 1-Jul-20 | 31-Jul-20 | 12248 | 0.22 | 1.00 | 1.00 | 0.01 | 0.96 | 0.96 | 1 |
| India | Kumar et al.<sup>46</sup> | 8-Oct-20 | 15-Oct-20 | 6366 | 0.32 | 1.00 | 1.00 | 0.02 | 0.92 | 0.94 | 2 |
| India | Sharma et al.<sup>47</sup> | 1-Aug-20 | 7-Aug-20 | 15046 | 0.28 | 0.92 | 0.98 | 0.04 | 0.95 | 0.99 | 4 |
| India | Sharma et al.<sup>47</sup> | 1-Oct-20 | 7-Oct-20 | 15015 | 0.26 | 0.99 | 0.99 | 0.04 | 0.96 | 0.997 | 5 |
| India | Sharma et al.<sup>47</sup> | 1-Sep-20 | 7-Sep-20 | 17409 | 0.25 | 0.99 | 0.99 | 0.09 | 0.90 | 0.98 | 10 |
| India | Kumar et al.<sup>48</sup> | 30-Sep-20 | 1-Oct-20 | 2000 | 0.08 | 0.93 | 1.00 | 0.09 | 0.74 | 0.82 | 10 |
| India | Khan et al.<sup>49</sup> | 17-Oct-20 | 4-Nov-20 | 6230 | 0.39 | 1.00 | 1.00 | 0.06 | 0.90 | 0.96 | 6 |
| Iran | Poustchi et al.<sup>50</sup> | 17-Apr-20 | 2-Jun-20 | 7745 | 0.15 | 0.72 | 0.98 | 0.45 | 0.36 | 0.6 | 70 |
| Iran | Shadmani et al.<sup>51</sup> | 15-Nov-20 | 20-Dec-20 | 1967 | 0.2 | 0.94 | 0.98 | 0.14 | 0.65 | 0.73 | 13 |
| Ireland | Heavey et al.<sup>52</sup> | 22-Jun-20 | 16-Jul-20 | 1732 | 0.02 | 0.94 | 1.00 | 0.26 | 0.27 | 0.36 | 33 |
| Italy | Melotti et al.<sup>53</sup> | 26-May-20 | 8-Jun-20 | 1813 | 0.26 | 1.00 | 1.00 | 0.46 | 0.17 | 0.3 | 81 |
| Italy | Pagani et al.<sup>54</sup> | 18-May-20 | 7-Jun-20 | 4143 | 0.22 | 0.97 | 0.96 | 0.27 | 0.31 | 0.35 | 17 |
| Kenya | Ngere et al.<sup>55</sup> | 2-Nov-20 | 23-Nov-20 | 1006 | 0.33 | 0.94 | 1.00 | 0.32 | 0.61 | 0.89 | 46 |
| Maldives | Abdul-Raheem et al.<sup>56</sup> | 15-Oct-20 | 16-Nov-20 | 1883 | 0.13 | 0.90 | 1.00 | 0.43 | 0.38 | 0.65 | 72 |
| Mali | Sagara et al.<sup>57</sup> | 28-Jul-20 | 16-Oct-20 | 2659 | 0.07 | 0.74 | 0.99 | 0.10 | 0.79 | 0.87 | 10 |
| Mexico | Basto-Abreu et al.<sup>58</sup> | 15-Aug-20 | 15-Nov-20 | 9464 | 0.23 | 0.92 | 1.00 | 0.08 | 0.67 | 0.72 | 8 |
| Mozambique | Arnaldo et al.<sup>59</sup> | 4-Aug-20 | 24-Aug-20 | 4719 | 0.04 | 0.87 | 0.99 | 0.15 | 0.71 | 0.76 | 9 |
| Mozambique | Arnaldo et al.<sup>59</sup> | 6-Jul-20 | 13-Jul-20 | 1358 | 0.03 | 0.83 | 0.98 | 0.16 | 0.77 | 0.81 | 7 |
| Mozambique | Arnaldo et al.<sup>59</sup> | 10-Aug-20 | 21-Aug-20 | 5066 | 0.03 | 0.83 | 0.98 | 0.04 | 0.88 | 0.83 | -5 |
| Netherlands | Vos et al.<sup>18</sup> | 31-Mar-20 | 11-May-20 | 3147 | 0.02 | 0.84 | 1.00 | 0.62 | 0.07 | 0.17 | 158 |
| Nigeria | Okpala et al.<sup>60</sup> | 8-Dec-20 | 15-Dec-20 | 3136 | 0.18 | 0.98 | 0.99 | 0.61 | 0.24 | 0.58 | 147 |
| Pakistan | Nisar et al.<sup>61</sup> | 15-Apr-20 | 22-Aug-20 | 3005 | 0.12 | 0.97 | 1.00 | 0.07 | 0.92 | 0.98 | 8 |
| Peru | Huamaní et al.<sup>62</sup> | 12-Sep-20 | 27-Sep-20 | 1920 | 0.33 | 0.97 | 1.00 | 0.55 | 0.30 | 0.65 | 119 |
| Peru | Díaz-Vélez et al.<sup>63</sup> | 24-Jun-20 | 10-Jul-20 | 2010 | 0.29 | 0.67 | 0.97 | 0.18 | 0.68 | 0.76 | 13 |
| Peru | Reyes-Vega et al.<sup>64</sup> | 28-Jun-20 | 9-Jul-20 | 3195 | 0.22 | 0.97 | 0.96 | 0.14 | 0.56 | 0.6 | 8 |
| Peru | Moyano et al.<sup>65</sup> | 11-Nov-20 | 30-Nov-20 | 1391 | 0.26 | 0.99 | 0.99 | 0.33 | 0.48 | 0.71 | 48 |
| Portugal | Canto e Castro et al.<sup>22</sup> | 8-Sep-20 | 14-Oct-20 | 13398 | 0.02 | 0.98 | 1.00 | 0.50 | 0.16 | 0.32 | 100 |
| Portugal | Kislaya et al.<sup>66</sup> | 21-May-20 | 8-Jul-20 | 2231 | 0.03 | 0.96 | 0.99 | 0.35 | 0.43 | 0.6 | 43 |
| Senegal | Talla et al.<sup>67</sup> | 25-Oct-20 | 26-Nov-20 | 1422 | 0.28 | 0.81 | 0.99 | 0.67 | 0.32 | 0.93 | 197 |
| Spain | Pérez-Gómez et al.<sup>68</sup> | 27-Apr-20 | 22-Jun-20 | 61092 | 0.04 | 0.91 | 0.99 | 0.33 | 0.29 | 0.42 | 48 |
| Switzerland | Richard et al.<sup>69</sup> | 6-Apr-20 | 30-Jun-20 | 8344 | 0.07 | 0.93 | 1.00 | 0.58 | 0.13 | 0.3 | 134 |
| UAE | Alsuwaidi et al.<sup>70</sup> | 19-Jul-20 | 14-Aug-20 | 8831 | 0.11 | 0.97 | 0.99 | 0.05 | 0.91 | 0.95 | 5 |
| UK | Ward et al.<sup>71</sup> | 20-Jun-20 | 13-Jul-20 | 99908 | 0.06 | 0.84 | 0.99 | 0.16 | 0.32 | 0.33 | 3 |
| UK | Ward et al.<sup>71</sup> | 31-Jul-20 | 13-Aug-20 | 105829 | 0.05 | 0.84 | 0.99 | 0.14 | 0.33 | 0.33 | -1 |
| UK | Ward et al.<sup>71</sup> | 15-Aug-20 | 28-Sep-20 | 159367 | 0.04 | 0.84 | 0.99 | 0.16 | 0.32 | 0.32 | -1 |
| US | Sullivan et al.<sup>19</sup> | 9-Aug-20 | 8-Dec-20 | 4654 | 0.05 | 0.98 | 0.99 | 0.80 | 0.10 | 0.47 | 386 |
| US | Lamba et al.<sup>72</sup> | 15-Aug-20 | 15-Dec-20 | 983 | 0.03 | 0.92 | 1.00 | 0.55 | 0.24 | 0.47 | 108 |
| US | Chamberlain et al.<sup>20</sup> | 9-Aug-20 | 8-Dec-20 | 1370 | 0.07 | 0.92 | 1.00 | 0.80 | 0.11 | 0.5 | 372 |
| US | Pathela et al.<sup>73</sup> | 13-May-20 | 21-Jul-20 | 40880 | 0.24 | 0.98 | 0.99 | 0.54 | 0.21 | 0.45 | 114 |

**Fig. 1.**
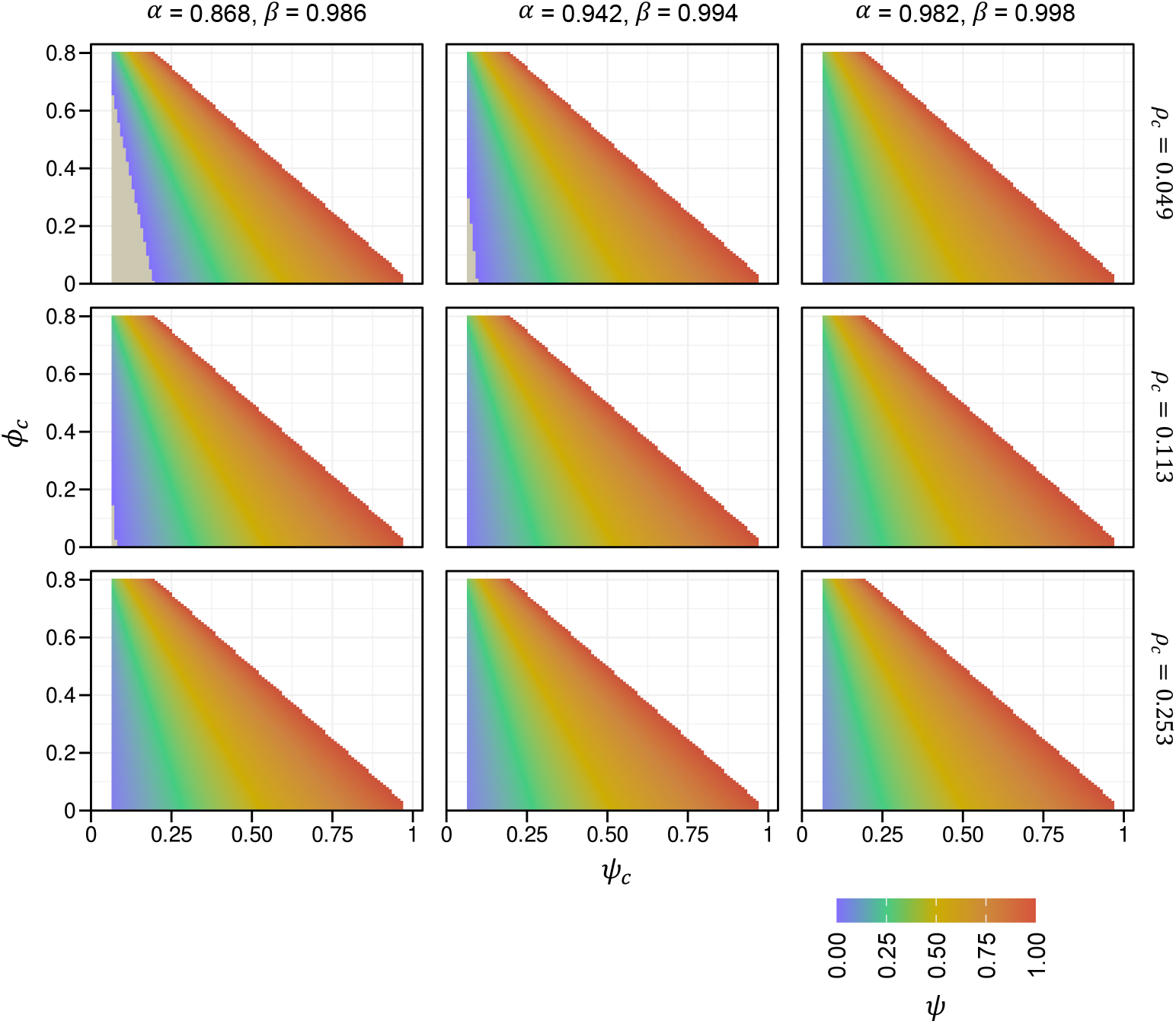
Dependence of proportion of asymptomatic infections on key survey characteristics. Estimates of *ψ* obtained using equation (1) for different combinations of the quantities in *Q* (see text) varied over ranges reflective of SARS-CoV-2 serosurveys (Table 1). Combinations of *α, β* and *ρ*_*c*_ used are indicated alongside each panel. Grey regions represent extreme parameter combinations (low values of several quantities in *Q*) for which equation (1) yields unrealistic estimates (*ψ* < 0).

Given the finite sample size, *N*, of a serosurvey, uncertainties bound the quantities in *Q*, which in turn would yield uncertainties in the estimates of *ψ*. To obtain the latter uncertainties, we developed a sampling procedure, which allowed the creation of multiple virtual instantiations of the serosurvey, each assuming distinct values of the quantities in *Q* drawn from their underlying distributions (Methods). Applying our formula to each of these instantiations yielded the corresponding distribution of *ψ* for the serosurvey. We tested our method against synthetic datasets next.

### Validation of formalism with synthetic serosurvey datasets

Generation of a synthetic dataset mimics a serosurvey *in silico*. It assumes known, or true, values of the parameters – *α, β, ρ, ϕ*, and *ψ* – defining a population, and then creates a sample of size *N* by subjecting each virtual individual in the sample to testing and symptom manifestation, simulating the results based on the parameters above (Methods). The resulting outcomes yield the quantities in *Q =* }*α, β, ρ*_*c*_, *ϕ*_*c*_, *ψ*_*c*_}, required for the application of our formalism. Our formalism would be deemed reliable if the resulting estimate of *ψ* were close to the value of *ψ* used to generate the synthetic dataset.

To test this, we generated a large number of synthetic datasets, spanning the range of parameter values seen in serosurveys (Table 1; Fig. 1). Thus, we chose the three combinations of *α* and *β* and the three values of *ρ* in Fig. 1. As a stringent test of our formalism, we chose broad distributions of *α* and *β* (setting *κ* = 30; see Text S2). Three values each of *ψ* and *ϕ* were chosen, to equal their first quartile, median, and third quartile from the serosurvey datasets (Table 1). Further, three sample sizes, *N*, were chosen, namely 1000, 5000, and 10000, again mimicking the ranges in the serosurveys. Together, these combinations yielded 243 synthetic serosurvey datasets. We applied our formalism to each of these datasets (Methods) and compared the estimated values of *ψ* with the true values used to generate the respective datasets. We performed the comparison by calculating Lin’s concordance correlation coefficient, which assesses the agreement between the two quantities in terms of accuracy, by determining how close the best-fit line is to the y=x line, and precision, by determining how close the points are to the best-fit line.^27^ The coefficient assumes values from -1 to 1, with 1 reflecting perfect concordance and -1 perfect inverse concordance. We found that our formalism accurately recovered the true values (Lin’s coefficient = 0.94; 95% CI, 0.92–0.95), indicating its reliability and robustness (Fig. 2). This gave us confidence in our formalism. We applied it next to SARS-CoV-2 serosurveys.

**Fig. 2.**
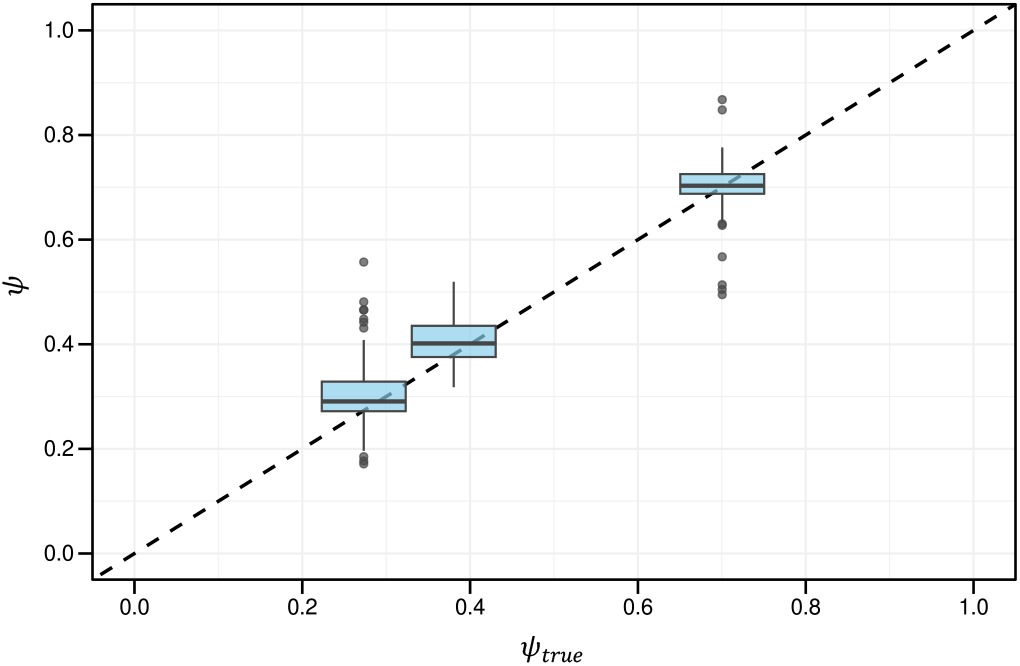
Validation using synthetic data. Comparison between *ψ* estimated using our framework applied to synthetic data and the true asymptomatic proportion (*ψ*_*true*_) used to generate the synthetic data (see Text). 243 synthetic datasets were generated by varying the sample size and the parameters *ρ, ψ, ϕ, α* and *β* over the ranges in Fig. 1. Box plots show the distribution of the estimated values of *ψ* for the three values of *ψ* chosen as inputs (*ψ*_*true*_). The box boundaries show quartiles, the central horizontal lines medians, whiskers represent 1.5 times the interquartile range beyond the box, and symbols are outliers. The dashed line is y=x. Lin’s coefficient = 0.94; 95% CI, 0.92– 0.95. The median absolute difference between *ψ* and *ψ*_*true*_ is 6.5%.

### Evidence of the confounding effect of symptom overlap in SARS-CoV-2 serosurveys

We collated data from published serosurveys, which rely on antibody-based tests for detecting SARS-CoV-2 infection and a questionnaire for assessing symptoms (Methods).^18-20,22,28-73^ Our method is applicable to surveys using nucleic acid-based (PCR) tests as well. Serosurveys have been preferred, however, for assessing asymptomatic SARS-CoV-2 infections because nucleic acid-based testing could miss presymptomatic individuals, who do not display symptoms at the time of testing but develop them later.^16,74^ Serosurveys seek symptoms experienced during a longer ‘recall period’, which renders them less likely to miss presymptomatic individuals. The longer recall periods, however, increase the chances of the tested individuals contracting other infections/conditions with overlapping symptoms during the recall periods, making serosurveys vulnerable to misclassification from symptom overlap and highlighting the need for our formalism.

We identified 50 serosurveys that satisfied our inclusion criteria (Methods).^18-20,22,28-73^ 15 (30%) of them were nation-wide serosurveys, 15 (30%) were conducted at subnational levels and 20 (40%) at local levels (Table S1). Four studies^31,47,59,71^ estimated *ψ*_*c*_ at three different time points, resulting in a total of 58 estimates of *ψ*_*c*_. We summarize the data collated in Table 1. The selected studies spanned 29 nations across Asia, the Americas, Europe, and Africa, covering a broad spectrum of epidemiological settings. Together, the studies sampled 847665 individuals. The crude seroprevalence, *ρ*_*c*_, varied from 0.01 to 0.58 across the studies, with a median of 0.11 (interquartile range: (0.049, 0.23)), implying varied extents of the spread of the infection in the populations studied at the time of the surveys (Fig. S2).

To assess the prevalence and scale of the effect of symptom overlap, we examined the fraction, *ϕ*_*c*_, of test-negative individuals who reported symptoms across the surveys. *ϕ*_*c*_ varied from 0 to 0.8 with a median of 0.28 (Table 1, Fig. 3a), indicating that overlapping symptoms commonly arose from other conditions and could therefore significantly affect estimates of *ψ*. Furthermore, although most surveys employed antibody tests with high sensitivity and specificity, several reported sensitivities ≤0.85 (Table 1), potentially amplifying the effect.

**Fig. 3.**
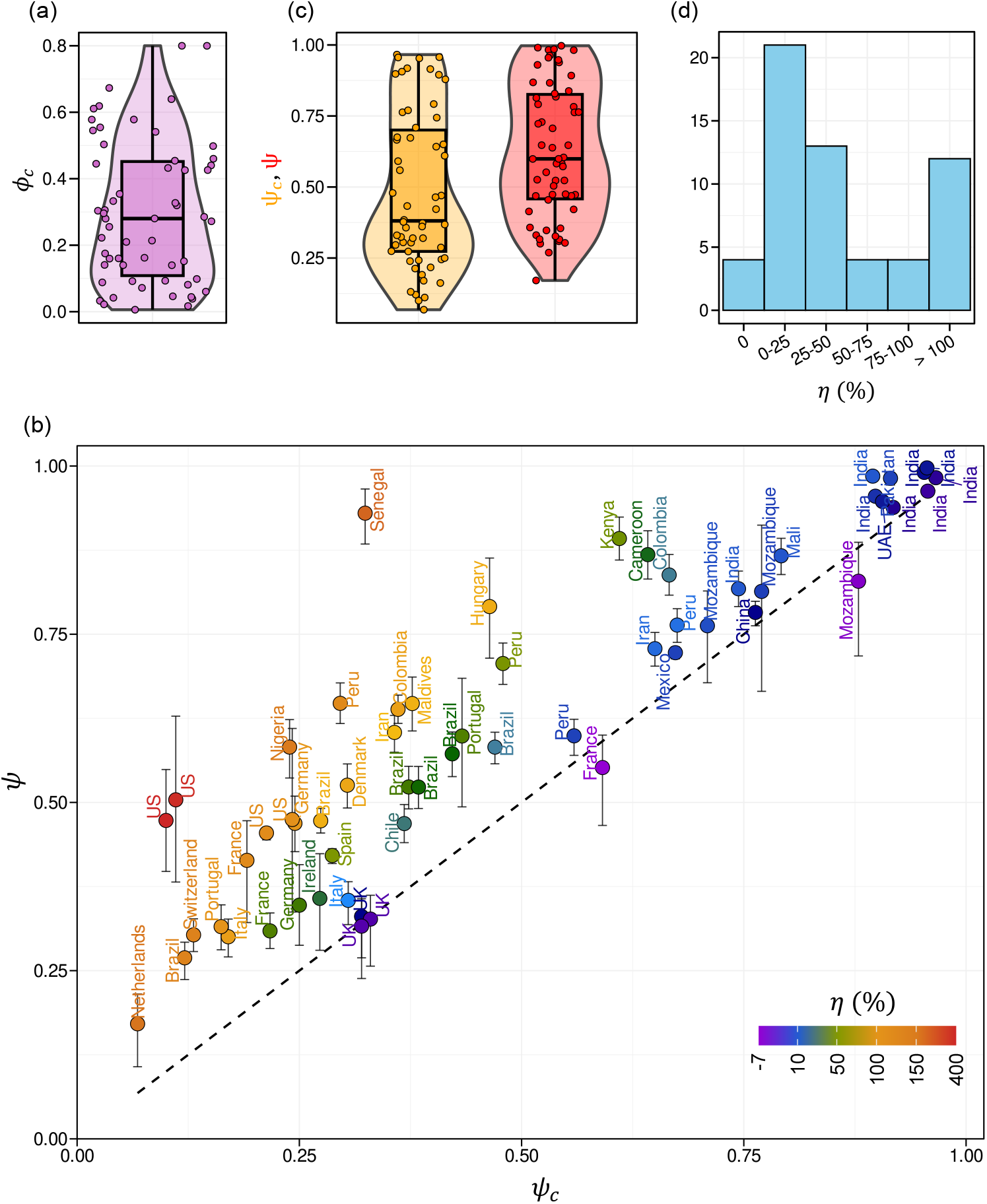
Accurate estimates of the proportion of asymptomatic infections. (a) The distribution (violin plot) of the estimates of *ϕ*_*c*_ (symbols), the fraction of test-negative individuals displaying symptoms, from 50 serosurveys (Table 1), offering evidence of symptom overlap. (b) Estimates of the asymptomatic proportion, *ψ*, obtained using our formalism (Methods), plotted against the reported estimates, *ψ*_*c*_, from the 50 serosurveys (Table 1). The country in which each serosurvey was conducted is indicated. Symbols are median values and error bars indicate interquartile ranges. The color coding is determined by the median *η =* 100 × (*ψ* − *ψ*_*c*_)*/ψ*_*c*_, shown in the scale bar. The dashed line represents *ψ = ψ*_*c*_. (c) Distributions of *ψ*_*c*_ (yellow) and *ψ* (red) for the 50 serosurveys. In (a) and (c), symbols are individual estimates (Table 1); widths of violins are proportional to the densities of the estimates; box plots depict quartiles, whiskers extreme values and horizontal lines medians. (d) Histogram showing the number of estimates in each bin of values of median *η* indicated.

### Estimates of the proportion of asymptomatic infections

The surveys reported widely varying estimates of the crude proportion of asymptomatic infections, *ψ*_*c*_, spanning the range from 0.07 to 0.97 (Table 1). Applying our formalism, we calculated the corrected proportion, *ψ*, for all the 58 estimates of *ψ*_*c*_. We found that *ψ* varied from 0.17 (IQR: (0.11, 0.22)) to 0.997 (IQR: (0.995, 0.999)) across the studies (Table 1, Table S2). Interestingly, we found that the median values of *ψ* thus estimated matched closely with the corresponding point estimates obtained using equation (1) (Table S2), indicating the utility of equation (1) when uncertainties in the quantities in *Q* are not available. Here, for greater reliability, we employed the median values of *ψ* and their interquartile ranges (IQR) for further analysis. Importantly, in 54 of the 58 cases, we found that *ψ* ≥ *ψ*_*c*_ (Fig. 3b). Indeed, *ψ* was significantly higher on average than *ψ*_*c*_; median *ψ*∼0.60 vs. median *ψ*_*c*_∼0.38 (P=8×10^-11^ using the Wilcoxon signed rank test) (Fig. 3c).

We defined *η =* 10000 × (*ψ* − *ψ*_*c*_)/*ψ*_*c*_ as the percentage increase in the present estimate over that reported. The symbols in Fig. 3b are color coded by their respective median estimates of *η* (accounting for the uncertainties in *ψ*; see Methods). Of the 58 estimates, 12 had *η*≥100%, 8 had *η* in the range of 50-100%, 13 in the range 25-50%, 21 between 0% and 25%, and 4 had *η*<0% (Fig. 3d). The highest were two studies from the US^19,20^ (387% (IQR: (362%, 401%) and 372% (IQR: (329%, 399%)), followed by Senegal^67^ (197% IQR: (188%, 206%)) and The Netherlands^18^ (158% IQR: (150%, 163%)) (Table 1). These large values of *η* imply that the proportion of asymptomatic infections was severely underestimated in these studies. Of the few instances where *ψ* was overestimated–specifically, the 4 studies with *η*<0%–the highest overestimation appeared in a study from France^39^ (-7% IQR: (-21%, 2%)) (Table 1). Nonetheless, the extents to which the present estimates deviate from the previously reported ones appear substantial across the studies, indicating the need to reassess the role of asymptomatic infections in the COVID-19 pandemic. Next, we examined the factors responsible for the deviations.

### Factors contributing to the correction in the estimate of the proportion of asymptomatic infections

From equation (1), we recognized that the quantities *Q =* }*α, β, ρ*_*c*_, *ϕ*_*c*_, *ψ*_*c*_} influence *ψ* and hence *η*. To assess the relative contributions of these quantities to the variation in *η* in the serosurveys we studied, we estimated the pairwise correlations between the quantities and *η*. While *α* showed no correlation, *β* showed a modest positive correlation with *η* (Spearman’s coefficient *r*_*S*_ = 0.28, P = 0.03) (Fig. S3). We found no correlation between the seroprevalence, *ρ*_*c*_, and *η* (Fig. S3), implying that the correction in the estimate of *ψ* due to our formula was not affected by the extent of spread of the infection in the population. *η*, however, was strongly positively correlated with *ϕ*_*c*_ (*r*_*S*_ = 0.96, P < 10^-16^) (Fig. 4).

**Fig. 4.**
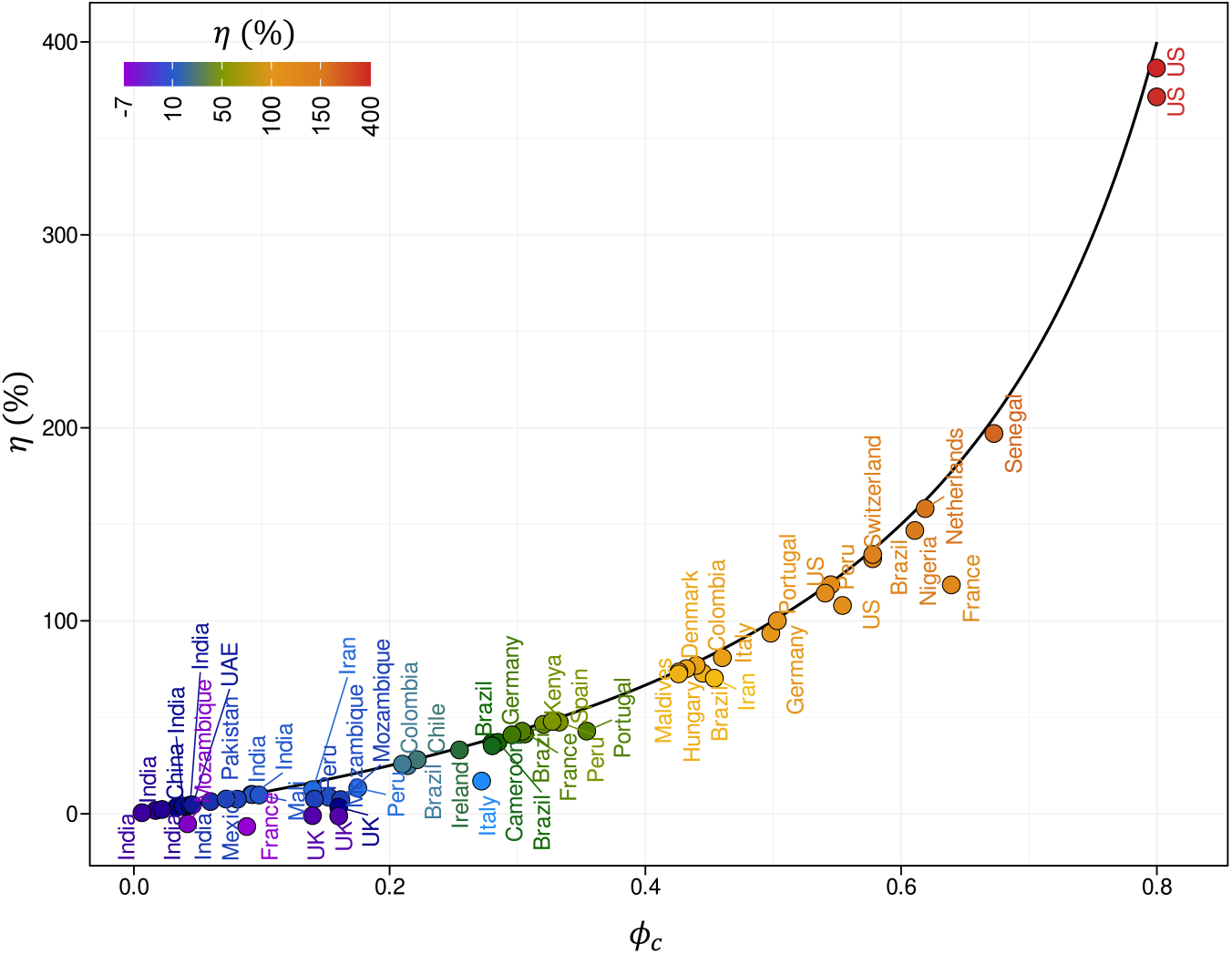
Dependence of the proportion of asymptomatic infections on the extent of symptom overlap. The relative increase in the proportion of asymptomatic infections estimated by our formalism, *η =* 100 × (*ψ* − *ψ*_*c*_)*/ψ*_*c*_, as a function of the fraction of test-negative individuals reporting symptoms, *ϕ*_*c*_, in the 50 serosurveys studied (symbols) (Table 1). The symbols are color-coded by the value of *η* (scale bar). *η* and *ϕ*_*c*_ are positively correlated (Spearman’s correlation, *r*_*s*_=0.96, P<10^-16^). The line is the median of 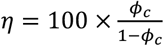, predicted using equation (2) (Methods). The prediction captures the data well (median *R*^*2*^=89.4%; IQR: (81.2%, 94.0%)).

To understand this latter correlation, because most serosurveys employed tests with high sensitivity and specificity, we considered the hypothetical scenario of the perfect test, where *α = β =* 1. In this scenario, equation (1) simplified to 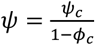, which yielded

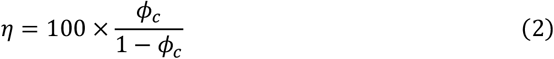

Thus, *η*>0 and increases monotonically with *ϕ*_*c*_, explaining the positive correlation between *η* and *ϕ*_*c*_. Indeed, the two studies from the US with the highest *η*^19,20^ both had *ϕ*_*c*_∼80%, the highest among all the surveys, and near perfect test sensitivity and specificity, explaining the large *η*. Note that the resulting estimates of *ψ*, 0.47 (IQR: (0.40, 0.55)) and 0.53 (IQR: (0.38, 0.63)), respectively, were consistent with independent estimates of *ψ*, namely, 0.47 (IQR: (0.34, 0.61)) and 0.45 (IQR: (0.44, 0.46)), obtained from two other surveys in the US^72,73^ where *ϕ*_*c*_ was much smaller (Table 1), indicating the reliability of the estimates offered by our formalism.

Remarkably, equation (2) closely recapitulated the dependence of *η* on *ϕ*_*c*_ from the 50 serosurveys, explaining 89.4% (IQR: (81.2%, 94.0%)) of the variation in *η* across the surveys (Fig. 4; Methods). The remaining 10.6% (IQR: (6%, 18.8%)) may be attributed to the other quantities in *Q* and any extraneous factors not included in our formalism.

To understand deviations from equation (2) as well as cases where *η*<0, we considered imperfect antibody tests, where *α* < 1 and/or *β* < 1. *ψ* then displayed a more complex dependence on the quantities in *Q* (equation (1)). In the absence of symptom overlap (*ϕ*_*c*_ *=* 0), equation (1) reduced to 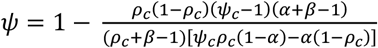, allowing *ψ* to be larger or smaller than *ψ*_*c*_ depending on the specific values of *α, β*, and *ρ*_*c*_. When *α =* 1, for instance, 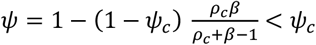. (The latter inequality follows because (1 − *β*)(1 − *ρ*_c_ ) > 0 and hence 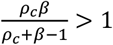.) Indeed, the reduction in *ψ* due to imperfect test sensitivity and specificity may dominate its increase due to overlapping symptoms, explaining the few instances with *η*<0 above. Nonetheless, in all but 4 of the 58 instances we studied, we found *ψ* ≥ *ψ*_*c*_, highlighting the dominant effect of symptom overlap. (We note that in no instance *ψ* > 1 (Table 1). It follows from equation (1) that *ψ* ≤ 1 so long as *ρ* > 0 (positive adjusted seroprevalence), *α* > *ρ*_*c*_ (sufficiently high test sensitivity), and *ψ*_*c*_ < 1 − *ϕ*_*c*_ (higher asymptomatic proportion in test-negative than test-positive individuals), all of which are typically expected. In the same way, no instances yielded *ψ* < 0, as no surveys had parameters belonging to the extreme conditions yielding these unrealistic estimates identified in Fig. 1.)

### Influence of potential confounding factors

We next examined whether factors beyond those in *Q* influenced our estimates of *ψ*, potentially confounding our inferences. We considered first the effect of sample size. We recall that the estimates of *ψ* and the associated uncertainties are obtained by sampling the quantities in *Q* as many times as the sample sizes in the respective surveys (Methods). If the sample sizes were inadequate to yield reliable estimates of *ψ*, one would expect an increase in the uncertainty in *ψ* with decreasing sample size. We found no significant correlation between the interquartile ranges of *ψ* estimated and the sample sizes from the serosurveys (Spearman’s coefficient=−0.17, P=0.2) (Fig. S4), ruling out sample size as a confounding factor.

Another factor affecting estimates of *ψ* may be the duration of the symptom recall period employed by the surveys. While symptoms typically last a short duration (∼days), antibodies are detectable in sera of infected individuals much longer (∼months) after infection.^75^ Thus, it is possible that individuals may be infected with symptoms before the recall period but have their symptoms resolved while still retaining antibodies during the recall period. Such individuals would be misclassified as asymptomatic, resulting in inflated estimates of *ψ*. If this effect were significant, we would expect a negative correlation of *ψ* with the recall period. For the surveys we examined, we found no significant correlation (Fig. S5; Spearman’s coefficient=0.18, P=0.17), implying that the variability in the recall period was not a significant factor affecting our estimates of *ψ*.

Similarly, heterogeneity in the symptoms assessed in the questionnaires across serosurveys may affect estimates of *ψ*. Specifically, surveys that assess more symptoms may identify more individuals in the uninfected compartment as symptomatic, as the symptoms may arise from a more diverse set of background conditions. To assess this possibility, we examined whether *ψ* was correlated with the number of symptoms assessed. 43 of the 50 surveys we studied provided explicit information of the symptoms. The number of symptoms varied substantially across surveys (median: 9; interquartile range: 6-11). All surveys included fever and all but one included cough (Fig. S6). We found no significant association between *ψ* and the number of symptoms (Spearman’s coefficient=0.06, P=0.70; Fig. S7a). To ensure that this lack of association was not an artefact of our formalism, we examined the association between the number of symptoms and the crude proportion of symptomatic individuals in the test-negative group, *ϕ*_*c*_. We again found no significant association (Spearman’s coefficient=0.25, P=0.09; Fig. S7b), suggesting that heterogeneity in symptom assessment across studies was not a key factor affecting our estimates of *ψ*. Our estimates of *ψ* were thus robust to these potential confounding factors.

### Improved consistency between estimates of the proportion of asymptomatic infections within nations

Following the improved consistency between the estimates of *ψ* in the US above, we asked whether a similar improvement would occur with other nations. More than one serosurveys were available for 12 nations (Table 1). We calculated the variance in the estimates of *ψ* and of *ψ*_*c*_, denoted *Var*(*ψ*) and *Var*(*ψ*_*c*_), respectively, across studies from individual nations (Table 2). Remarkably, we found that *Var*(*ψ*) was significantly lower than *Var*(*ψ*_*c*_) overall (P=0.014 using a 1-tailed paired Student’s t-test) (Table 2). The highest reduction, of 92%, in the variance due to our formalism was for the US. The next highest reduction was for Italy and Mozambique (84% each), followed by Iran (82%) (Table 2). For two nations, namely, Germany and the UK, *Var*(*ψ*) was larger than *Var*(*ψ*_*c*_). This was because in both these cases, *Var*(*ψ*_*c*_) *≈* 0, which may be coincidental given that they had low test sensitivities (<90%) in at least one study each (Table 1). Thus, our formalism reduced the heterogeneity between the estimates of *ψ* obtained from different studies in the same nation.

**Table 2.** Improved consistency in estimates of proportion of asymptomatic infections. The variance in *ψ*_*c*_ and *ψ* and their relative difference across multiple studies (estimates) from individual nations. The studies used for each country listed are in Table 1.

| Country | Number of studies | $Var(\psi_c)$ | $Var(\psi)$ | Reduction<sup>\$</sup> (%) |
| --- | --- | --- | --- | --- |
| US | 4 | 0.005 | 0.0004 | 92 |
| Italy | 2 | 0.009 | 0.001 | 84 |
| Mozambique | 3 | 0.007 | 0.001 | 84 |
| Iran | 2 | 0.043 | 0.008 | 82 |
| Peru | 4 | 0.025 | 0.005 | 80 |
| France | 3 | 0.050 | 0.015 | 70 |
| Colombia | 2 | 0.047 | 0.020 | 57 |
| India | 8 | 0.005 | 0.003 | 36 |
| Brazil | 6 | 0.016 | 0.013 | 16 |
| Portugal | 2 | 0.037 | 0.040 | -9 |
| UK* | 3 | 0.000 | 0.0001 | — |
| Germany* | 2 | 0.000 | 0.007 | — |

<sup>\$</sup>Calculated as $\frac{Var(\psi_c) - Var(\psi)}{Var(\psi_c)} \times 100$
\*Reduction is indeterminate because $Var(\psi_c) \approx 0$

Interestingly, when we considered all the 58 estimates together, *Var*(*ψ*) was lower than *Var*(*ψ*_*c*_) by 26%, but this difference did not reach statistical significance (P=0.29 using Levene’s test). This implied that variations in *ψ* across nations introduced by other factors, such as racial, ethnic and demographic differences, which are yet to be fully understood, outweighed the variations introduced by overlapping symptoms and test sensitivity and specificity, the latter accounted for in our formalism.

### Improved association of the proportion of asymptomatic infections with age

Several studies have highlighted the increasing occurrence of symptoms with age.^76,77^ To examine this dependence, we collated the median ages of the samples in the serosurveys we studied (Table S1) and estimated their association with *ψ* and *ψ*_*c*_.

Expectedly, we found both *ψ* and *ψ*_*c*_ to be strongly negatively associated with age (Fig. 5), consistent with previous studies.^76,77^ Interestingly, the association was much stronger with *ψ* (Spearman’s coefficient=-0.69, P=2×10^-9^) than with *ψ*_*c*_ (Spearman’s coefficient=-0.60, P=8×10^-7^). Thus, correcting for the uncertainties introduced by symptom overlap and test sensitivity and specificity revealed a much greater dependence of *ψ* on age than previously estimated, reiterating the advantage of our formalism in better characterizing epidemiological and demographic underpinnings of asymptomatic infections.

**Fig. 5.**
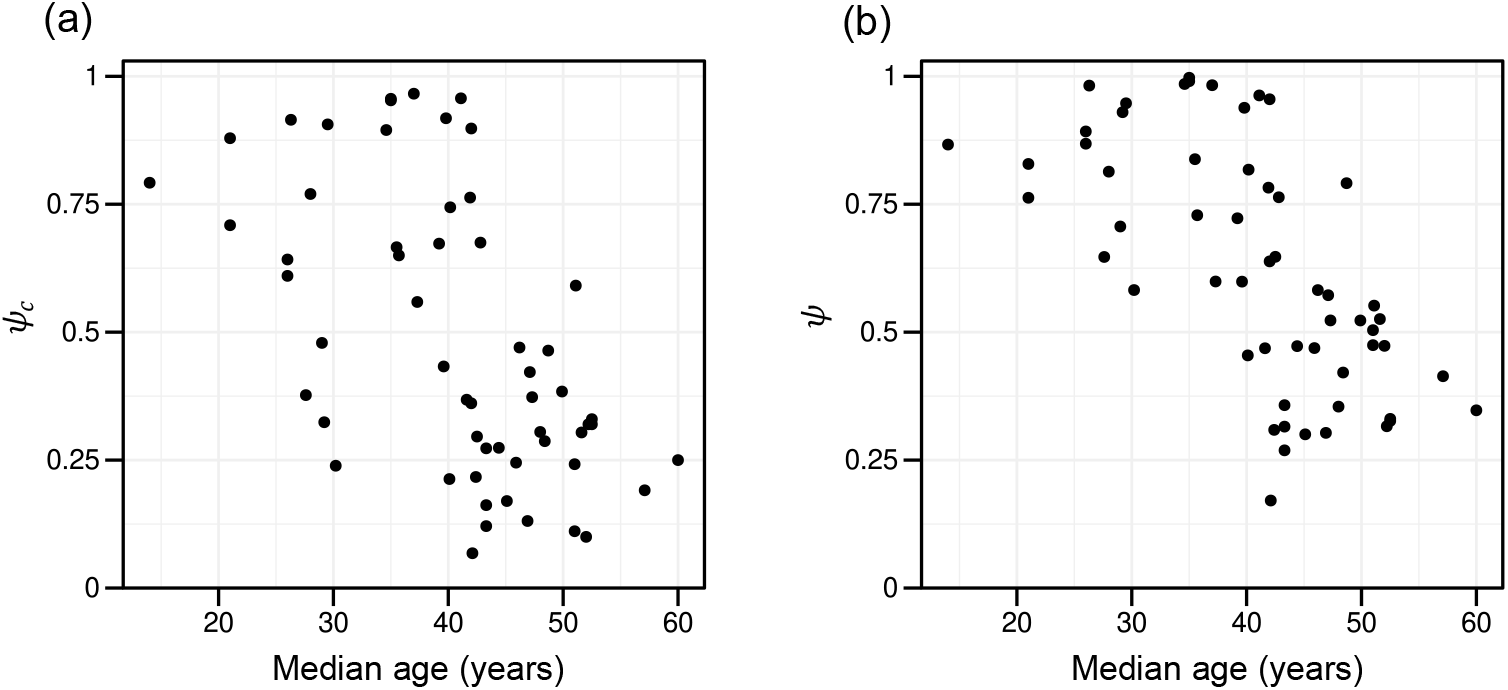
Association between age and proportion of asymptomatic infections. Association between median age of study participants in serosurveys (Table S1) and the corresponding estimate of (a) *ψ*_*c*_ (Spearman’s coefficient = -0.60, P = 8×10^-7^) and (b) *ψ* (Spearman’s coefficient = -0.69, P = 2×10^-9^)

### Validation against PCR test-based studies

To validate our formalism further, going beyond synthetic datasets, we sought direct comparisons with independent, PCR-based studies. The latter studies offer an alternative route to serosurveys for estimating *ψ*. We recall that a key limitation of these studies is their tendency to miss presymptomatics. If such studies employed a >2-week follow-up to identify presymptomatics, they would yield reliable estimates of *ψ*,^74^ offering a test of our formalism.

As a stringent test of our formalism, we first looked for surveys that performed PCR as well as serology tests on the same sample. After a comprehensive search (Methods), we identified a study on the staff and residents of long-term care facilities (LTCFs) in New Jersey, US performed in the early days of the pandemic.^78^ PCR test results were available for all staff and residents by May 19, 2020, as per the directive of the state. Between October 2020 and March 2021, serology and symptomatology tests were performed on the staff and residents of 10 LTCFs participating in the study. Because of the small sample size of the residents and their advanced age, we considered data on the staff. For 299 individuals, valid RT-PCR, antibody, and symptom data were available, allowing a direct test of our formalism. The crude estimates from the PCR and serology data were *ψ*_*c*_=0.36 and *ψ*_*c*_=0.41, respectively. Applying our formalism to the data yielded the corresponding estimates of *ψ*=0.43 (IQR:(0.38, 0.47)) and *ψ*=0.46 (IQR:(0.42, 0.50)), respectively, bringing the estimates closer to each other than the crude estimates (Table 3). Because the symptom recall period was large, some PCR-negative individuals also reported symptoms, reiterating the confounding effect of symptom overlap. The resulting improved agreement between PCR-based and serology-based estimates of *ψ* supported the application of our formalism.

**Table 3.** Comparison between PCR- and antibody test-based estimates. Symbols have the same meaning as in Table 1.

| Comparison | Survey | Study period | N | $\rho_c$ | $\alpha$<br>(95% CI) | $\beta$<br>(95%CI) | $\phi_c$ | $\psi_c$ | $\psi$<br>(IQR) |
| --- | --- | --- | --- | --- | --- | --- | --- | --- | --- |
| Same sample<sup>78</sup> | PCR-based* | 19-May-20 | 299 | 0.27 | 0.87<br>(0.81, 0.91) | 1<br>(0.96, 1.00) | 0.20 | 0.36 | 0.43<br>(0.38, 0.47) |
|  | Antibody-based | 20-Oct-20 to 04-Mar-21 | 299 | 0.39 | 0.98<br>(0.90, 1.00) | 0.99<br>(0.98, 1.00) | 0.14 | 0.41 | 0.46<br>(0.42, 0.50) |
| Distinct samples | PCR-based<sup>79</sup> | 25-Apr-20 to 29-Apr-20 | 3605 | 0.02 | 0.87<br>(0.81, 0.91) | 1<br>(0.96, 1.00) | 0<sup>†</sup> | 0.44 | 0.37<br>(0.27, 0.44) |
|  | Antibody-based<sup>19</sup> | 09-Aug-20 to 08-Dec-20 | 4654 | 0.05 | 0.98<br>(0.90,1.00) | 0.99<br>(0.98, 1.00) | 0.80 | 0.10 | 0.47<br>(0.40, 0.55) |
|  | Antibody-based<sup>73</sup> | 13-May-20 to 21-Jul-20 | 40880 | 0.24 | 0.98<br>(0.96,0.99) | 0.99<br>(0.98,0.99) | 0.54 | 0.21 | 0.45<br>(0.44, 0.46) |

<sup>†</sup>Assumed based on the short, 2-week follow-up

To ascertain the reliability of our formalism further, we relaxed the stringency above and sought comparisons between PCR-based and serology-based surveys on similar but not identical samples. Thus, we sought PCR-based surveys on samples similar to those in our serosurvey studies above (Table 1).^18-20,22,28-73^ Again, from a comprehensive literature search (Methods), we identified a study in the US that satisfied our criteria.^79^ This study was conducted during April 25-29, 2020 on a state-wide random sample of adults in Indiana, US. It performed RT-PCR tests and sought symptoms experienced over 2 weeks prior to testing. This ensured that most asymptomatic individuals were identified, as symptoms typically appear within 2 weeks of infection. From a large sample of 3605 tested individuals, it reported *ψ*_*c*_=0.44. Data of *ϕ*_*c*_ was not reported. Recognizing that the chances of symptoms arising from an independent condition in two weeks are small, we rederived our expression above (equation (1)), obviating the need for estimates of *ϕ*_*c*_ (Methods). Applying the expression yielded *ψ*=0.37 (IQR:(0.27, 0.44)). In comparison, applying our formalism to data on 4654 individuals from a nationwide serosurvey of the US conducted during August-December, 2020^19^, which reported *ψ*_*c*_=0.1, yielded *ψ*=0.47 (IQR: (0.40, 0.55)). The wide confidence intervals were due to the low seroprevalence in the two studies (*ρ*_*c*_=0.02 and 0.05, respectively; Table 3). Notwithstanding, the much closer agreement between the two estimates of *ψ* is noteworthy given the wide disparity in the corresponding crude estimates (*ψ*_*c*_=0.44 and 0.1, respectively).

To avoid any confounding effects arising from the time-gap between the two studies, we examined a third study, a much larger serosurvey in the city of New York, conducted between May and July, 2020. The study reported *ψ*_*c*_=0.21. Our formalism applied to the data yielded *ψ*=0.45 (IQR:(0.44, 0.46)) (Table 3), again in far closer agreement with the other two estimates than the corresponding crude estimates, together offering yet another successful test of our formalism. These successful comparisons, both within and between samples, suggested that our formalism was reliable.

### Application to other pathogens: Zika virus serosurveys

Finally, to test the wider applicability of our formalism, we applied it to data from 3 Zika virus serosurveys. Specifically, the surveys were conducted during the Zika virus outbreaks in Yap (2007)^80^, French Polynesia (2013-2014)^81^, and Puerto Rico (2016)^82^, respectively. The surveys reported all the data required for us to apply our formalism (Table 4). The crude seroprevalence levels varied substantially across the studies (*ρ*_*c*_∼0.26–0.74). Importantly, they reported substantial fractions of test-negative individuals displaying symptoms (*ϕ*_*c*_∼0.13–0.27), reiterating the need to correct for symptom overlap in estimating *ψ*. Using our formalism, we found again that *ψ* was substantially higher than *ψ*_*c*_ in the Yap (median 0.73 vs. 0.62) and French Polynesia (median 0.56 vs. 0.43) surveys and remained unchanged in the Puerto Rico (both 0.51) survey (Table 4). Interestingly, a Bayesian inference framework, which also accounts for symptom overlap and test sensitivity and specificity, had been applied to these datasets earlier to estimate *ψ* and its uncertainties^7^, allowing us to compare our formalism with an independent approach. We found our estimates to be in close agreement with those from the latter (Table 4), yielding yet another validation of our formalism, and signifying its broad applicability, beyond SARS-CoV-2.

**Table 4.**
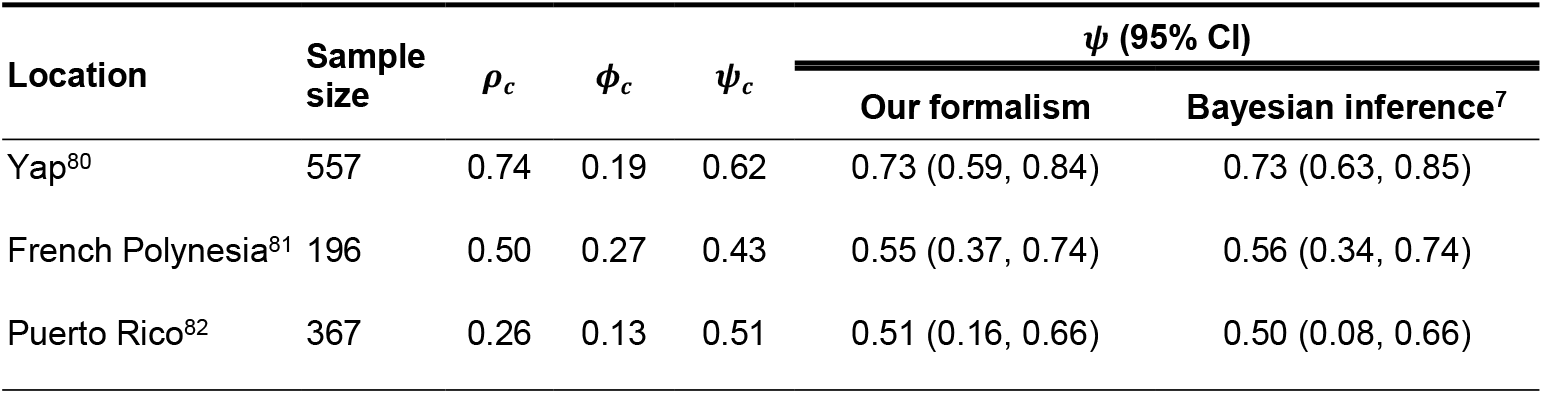
Application to Zika virus serosurveys. Data reported from three Zika virus serosurveys, indicated by their locations, were used to estimate the proportion of asymptomatic infections our formalism. Estimates reported from the Bayesian framework implemented previously^7^ are also shown. Symbols have the same meanings as in Eq. (1). Assay sensitivity and specificity were chosen according to the priors reported for all the three surveys^7^: *α*∼*Beta*(*35, 2*) and *β*∼*Beta*(1*7*, 1).

### Web application

We developed a web application, ShinyPsi, which enables the ready application of our formalism to serosurvey data. The URL for the application along with instructions for its usage is: https://therapeuticengglab.github.io/ShinyPsi/. The application runs locally, within the web browser, and needs no server host. No user data is collected, guaranteeing data protection. The application runs effortlessly on most machines, yielding results in seconds. We tested ShinyPsi on standard laptops with MacOS (browser: Safari, 16 GB RAM), Windows OS (browser: Chrome, 16GB RAM) and Linux (browser: Firefox, 8GB RAM). On these laptops, running the simulation with 10^5^ instantiations took approximately 10 seconds, 15 seconds and 30 seconds, respectively.

## DISCUSSION

In this study, we present a new formalism that employs data from population-based surveys and yields accurate estimates of *ψ* by accounting for assay sensitivity and specificity and the effect of symptom overlap with other conditions. We derived a closed-form, analytical expression for *ψ* (equation (1)) which enables ready application of our formalism. By analyzing data from 50 SARS-CoV-2 serosurveys, we found that asymptomatic infections are far more prevalent than previously estimated. Their role in the spread of the pandemic is thus likely to be far greater than previously anticipated. We applied our formalism to Zika virus serosurveys as well, highlighting its applicability beyond SARS-CoV-2. We developed a freely accessible web application that could be run locally, enabling the wide, safe, and easy implementation of our formalism.

Our formalism makes important conceptual and technical advances over prevalent methods^1,5-7,22,23^ to estimate *ψ*. The Rogan-Gladen estimator offered a closed-form analytical expression to estimate the seroprevalence by adjusting for test sensitivity and specificity.^21^ Despite several attempts^7,22,23^, a similar formula had not been forthcoming for *ψ*. This was because of the complexity involved in simultaneously correcting for the effects of test sensitivity and specificity as well as symptom overlap. Here, through rigorous mathematical reasoning, we overcame this challenge. Our derivation thus culminated in a closed-form expression for *ψ* (equation (1)).

The closed-form expression allows us to see directly how the key factors influencing *ψ* are related, offering quantitative insights into their effects. Our formalism showed, for instance, that symptom overlap was responsible for nearly 90% of the variation in the estimates of *ψ* across the 50 SARS-CoV-2 serosurveys we analyzed. The other factors involved, especially test sensitivity and specificity, seemed to have a more modest effect. Accordingly, we derived an even simpler expression correcting for symptom overlap alone (equation (2)), which could be used to obtain approximate estimates of *ψ* when test sensitivity and specificity are unknown (Fig. 4). When the latter estimates are available but not their distributions, as is often the case (Table S3), more refined approximations of *ψ* than offered by equation (2) can be obtained as point estimates from equation (1). The point estimates tend to be close to the median values of *ψ* obtained by accounting for the uncertainties in all the factors in our formalism (Table S2). Indeed, when the latter uncertainties are known, our formalism estimates the entire distribution of *ψ*, enabling its complete characterization.

The closed-form expression also makes our formalism computationally hugely efficient compared to existing methods. The only available alternative to our method is the Bayesian inference technique^7^. Like our formalism, that technique too starts with the basic equations for the joint probabilities of individuals in a serosurvey testing positive or negative and/or showing symptoms or not. It, however, does not perform any mathematical development beyond the basic equations and resorts to parameter estimation using Markov chain Monte Carlo (MCMC) sampling given the serosurvey data. One of the parameters thus estimated is *ψ*. This approach makes the technique computationally expensive. Each estimate of *ψ* for the Zika virus outbreaks, to which the technique was applied (Table 4), required running 3 independent MCMC chains with 1,000,000 iterations per chain.^7^ In contrast, our analytical formula offers a more sophisticated starting point for the estimation of *ψ*, which translates into huge gains in computational efficiency without compromising accuracy. Simple sampling of the parameters influencing *ψ* (equation (1)), often as few as ∼1000 times, yielded estimates of *ψ* along with their uncertainties that were in close agreement with the Bayesian technique (Table 4). This efficiency allowed us to test and apply our formalism at scale: we validated it against nearly 250 synthetic datasets, spanning wide parameter ranges, and applied it to 50 SARS-CoV-2 serosurveys. As we mention earlier, the Bayesian inference technique, in contrast, is yet to be applied to any SARS-CoV-2 dataset despite its development before the COVID-19 pandemic.

Because of the computational efficiency, we were able to create a live web application of our formalism that runs locally, with no requirement of a back-end server for computation, and yields estimates of *ψ* and its uncertainties in seconds. This ease of use is likely to enable wide application of our formalism. An added advantage of the locally run application is that it ensures complete data protection, with no exchange with any back-end service provider.

Insights from our study extend beyond the effects of test sensitivity and specificity and symptom overlap, which our formalism explicitly considered. Our findings revealed that estimates of *ψ* from different studies within the same nation tended to be more consistent than previously estimated. Interestingly, and in striking contrast, the disparity between estimates across nations did not get significantly reduced by the application of our formalism, suggesting that factors beyond those considered in our study were responsible for the variation across nations. Future studies may identify these factors. Similarly, our study showed that *ψ* was more strongly associated with age than previously estimated. At the same time, it ruled out sample size, symptom recall period, and heterogeneity in the symptoms considered across serosurveys as factors significantly affecting estimates of *ψ*. These insights not only help better design and interpret serosurveys, but also help sharpen the focus of the significant efforts underway to unravel genetic, immunological, and demographic underpinnings of asymptomatic SARS-CoV-2 infections.^83-87^

We foresee further implications of our study. The estimates of *ψ* that our formalism yields would help reassess the contribution of asymptomatic infections to COVID-19 transmission and spread.^8,12^ They would also form inputs to models of COVID-19 epidemiology^13-15^, enabling more reliable forecasting of disease spread and the design of effective control strategies. Similarly, the formalism could aid COVID-19 vaccine development efforts^88^ by enabling more accurate estimation of vaccine efficacies, which are often based on comparing estimates of *ψ* in the vaccinated and unvaccinated arms of clinical trials.^89-91^

We demonstrated the applicability of our formalism to settings beyond COVID-19. We showed how estimates of *ψ* could be obtained for Zika virus (Table 4). We anticipate the application of our formalism to other globally important pathogens that can cause asymptomatic infections.^1-4^ For instance, in a study estimating Ebola prevalence using highly sensitive and specific antibody assays, over 80% of symptomatic individuals were found to be seronegative, clearly indicating that the investigated symptoms were caused by other background infections.^3^ Tracking asymptomatic infections is critical for pathogens like Ebola due to their high mortality rate; asymptomatic individuals may act as silent drivers of transmission and the associated mortality. Identifying asymptomatic infections may be important independently of their role in preventing transmission. For example, asymptomatic malaria has been associated with chronic, low-grade hemolysis and intermittent, higher-density symptomatic recurrences.^4^ Identifying these individuals may help initiate treatments that may prevent these long-term adverse consequences of the infections. Our formalism may help anticipate the proportion of these individuals better and intensify screening efforts to identify them.

Our study has limitations. First, our formalism does not consider individuals infected by the pathogen of interest more than once but displaying symptoms only on some of those occasions. The chances of reinfection by SARS-CoV-2 were rare during the recall periods in the surveys we examined^92,93^, justifying our formalism. Yet, given that reinfections tend to be milder^92,93^, as also suggested by vaccination studies^89-91^, accounting for reinfections may further refine our estimates of *ψ*. Second, we assumed that symptoms caused by the pathogen of interest and by other infections/conditions are independent. While co-infection can potentially influence the severity of SARS-CoV-2 infection, such instances appear rare.^94,95^ Further justification of our assumption comes from studies that found influenza vaccination not to offer significant protection against SARS-CoV-2 symptoms.^96^ Whether similar justification can be found for other pathogens to which our formalism may be applied remains to be ascertained. Finally, our selection of SARS-CoV-2 serosurveys was not exhaustive. Our aim was to demonstrate the wide applicability and relevance of our formalism and not to provide a global estimate of *ψ* for SARS-CoV-2. Future studies may conduct a more systematic search and meta-analysis, using our formalism, to obtain a global estimate of *ψ*.

## METHODS

### Formalism to estimate the proportion of asymptomatic infections from surveys

We consider the scenario where infection by the pathogen of interest, denoted *X*, can trigger symptoms that may also be triggered by other pathogens (or conditions), the latter collectively denoted *Y*. Surveys aim to assess the proportion of asymptomatic infections by *X*. A test, denoted *T*, assesses whether an individual undertaking the test is infected by *X*. Simultaneously, a questionnaire inquires into the symptoms, denoted *S*, experienced by the individual during a pre-defined recall period. We recognize that the symptoms may also be triggered by *Y*. We distinguish between these possibilities by letting *S*_*X*_ and *S*_*Y*_ represent events associated with the symptoms being triggered by *X* and *Y*, respectively. The aim is to estimate the fraction of individuals infected by *X* who do not experience symptoms triggered by *X*. We arrive at this estimate as follows.

We define *P*[*X*^+^] and *P*[*T*^+^] as the probability with which an individual is infected by *X* and the probability that the infection test yields a positive result, respectively. Clearly, *P*[*T*^+^] *= ρ*_*c*_, the crude prevalence estimated by the survey as the fraction of individuals tested who show a positive result. *P*[*X*^+^] *= ρ* is the actual prevalence, obtained after correcting for test sensitivity and specificity. The test sensitivity is *α = P*[*T*^+^|*X*^+^], the probability of the test yielding a positive result given the infection by *X*. The test specificity is *β = P*[*T*^−^|*X*^−^], the probability that the test yields a negative result given that the tested individual is not infected by *X*. The total probability of the test yielding a positive result can thus be written as

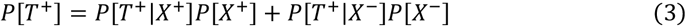

Recognizing that *P*[*T*^+^|*X*^−^] *=* 1 − *P*[*T*^−^|*X*^−^] and *P*[*X*^−^] *=* 1 − *P*[*X*^+^] and substituting the definitions above in equation (3), it follows that

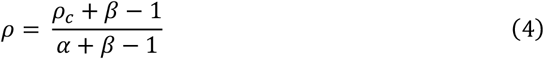

Note that equation (4) is the well-known Rogan-Gladen estimator of seroprevalence.^21^ We next consider events related to the occurrence of symptoms. The crude proportion of asymptomatic individuals, *ψ*_*c*_ *= P*[*S*^−^|*T*^+^], is the probability that an individual who tests positive reports no symptoms. It is thus measured in the surveys as the fraction of test-positive individuals who declare no symptoms. Accounting for the test sensitivity and specificity, we again write,

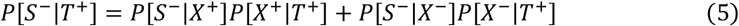

which, upon recognizing that *P*[*X*^−^|*T*^+^] *=* 1 − *P*[*X*^+^|*T*^+^] and invoking the identity involving conditional probabilities, *P*[*X*^+^|*T*^+^]*P*[*T*^+^] *= P*[*T*^+^|*X*^+^]*P*[*X*^+^], so that

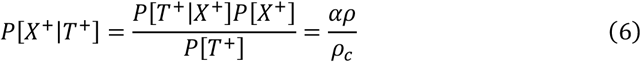

yields

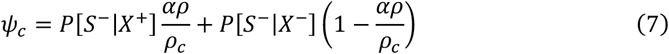

Given the simultaneous presence of *X* and *Y* in the population, the absence of symptoms implies the absence of symptoms triggered by both *X* and *Y*. In other words, }*S*^−^} *=* 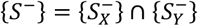. This yields,

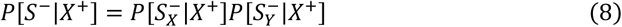

where 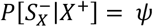 is the probability that an individual infected by *X* does not experience symptoms triggered by *X*, which is the true proportion of asymptomatic infections, the key quantity of interest here.

Similarly, in the absence of infection by *X*, we may write

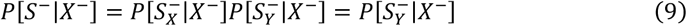

where the latter equality follows from 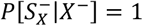; an individual not infected by *X* cannot have symptoms triggered by *X*.

Combining equations (7)-(9) yields

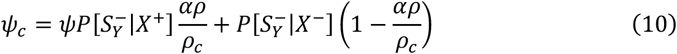

We next assume that experiencing symptoms triggered by *Y* is independent of infection by *X*, so that

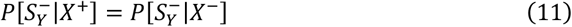

For SARS-CoV-2, this assumption follows from studies that found that influenza vaccination did not reduce COVID-19 symptoms.^96^ While co-infection with another respiratory pathogen can potentially influence the severity of SARS-CoV-2 infection, such instances appear rare, especially given the historically low prevalence of such pathogens during the early phases of the pandemic.^94,95^ To estimate the latter probabilities, we invoke their relationship with test results as follows. We recognize that *ω*_*c*_ *= P*[*S*^−^|*T*^−^] is the probability of not experiencing symptoms given test-negative status, which represents the crude proportion of asymptomatic individuals among test-negative individuals. Following the arguments above, the symptoms must arise neither from *X* nor from *Y*, so that

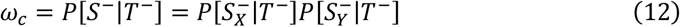

Invoking test sensitivity and specificity, we write the first term on the right-hand side of equation (12) as

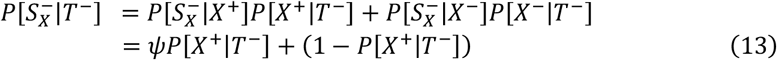

where the latter equality follows because 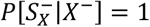 and *P*[*X*^−^|*T*^−^] *=* 1 − *P*[*X*^+^|*T*^−^]. Using the identity *P*[*X*^+^|*T*^−^]*P*[*T*^−^] *= P*[*T*^−^|*X*^+^]*P*[*X*^+^] and the definitions of the quantities above, we obtain

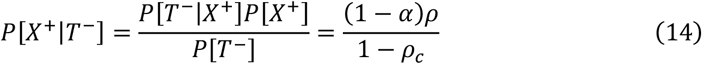

Combining equations (13) and (14) and rearranging terms yields

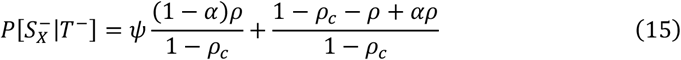

Following a similar procedure, we write the second term on the right-hand side of equation (12) as

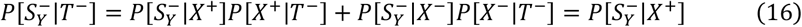

where the latter equality follows because 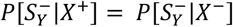 and *P*[*X*^+^|*T*^−^] *=* 1 − *P*[*X*^−^|*T*^−^]. Combining equations (15) and (16) with equation (12) and rearranging terms, we obtain

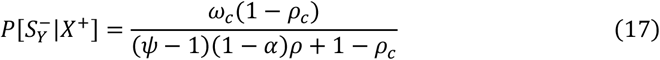

Finally, combining equations (10)-(12) and (17), and letting *ϕ*_*c*_ *=* 1 − *ω*_*c*_, the fraction of symptomatic individuals in the test-negative subpopulation, we obtain equation (1):

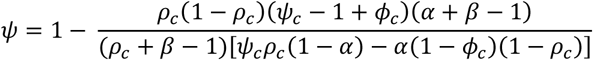

Equation (1) offers a readily applicable formula to obtain accurate estimates of the proportion of asymptomatic infections from survey data after accounting for assay sensitivity and specificity as well as symptom overlap with other conditions.

### Generation of synthetic datasets

Generation of a synthetic dataset involves the backward instantiation of a sample mimicking a serosurvey given the true features of the population. Thus, we assumed known (or true) values of the seroprevalence, *ρ*, the asymptomatic proportion, *ψ*, the proportion of uninfected individuals who displayed overlapping symptoms due to other conditions, *ϕ*, and distributions of test assay sensitivity and specificity, *α*∼*Beta*(*a, b*) and *β*∼*Beta*(_*c*_, *d*), respectively. Note that even with the same testing kits, sensitivity and specificity may vary across individuals, due to factors such as age, antibody levels, the diversity of antibody responses, severity of infection, and timing of measurement^91,97,98^, which are collectively accounted for in the latter distributions. We let the dataset contain *N* individuals, the sample size. We simulated data associated with the individuals as follows.

For each individual *j ∈* (1,, *… N*), we first identified the infection status as the outcome of a Bernoulli trial, *I*_*j*_∼*Bernoulli*(*ρ*), where success, *I*_*j*_ *=* 1, represents infection and failure, *I*_*j*_ *=* 0, implies otherwise. We next identified the outcome of testing, again as Bernoulli trials, conditional on infection status. For the *j*^*th*^ individual, we determined the assay sensitivity and specificity by sampling from the respective distributions: *α*_*j*_∼*Beta*(*a, b*) and *β*_*j*_∼*Beta*(_*c*_, *d*). Then, the test status was *T*_*j*_∼*Bernoulli*(*α*_*j*_) if *I*_*j*_ *=* 1; else, *T*_*j*_∼*Bernoulli*(1 − *β*_*j*_). Together, a positive test result was represented as *T*_*j*_ *=* 1 and a negative test result by *T*_*j*_ *=* 0. From this, the crude seroprevalence of the synthetic survey was obtained as

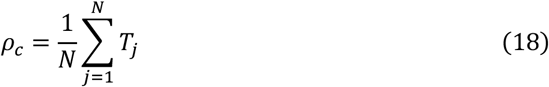

We next simulated the symptom status of each individual. If the *j*^*th*^ individual were infected, symptoms could arise from the infection and/or other conditions, whereas if the individual were uninfected, then symptoms could only arise from other conditions. We let *Sj* represent the symptom status of the *j*^*th*^ individual, with *S*_*j*_ *=* 1 implying symptomatic and *S*_*j*_ *=* 0 asymptomatic status. It followed that if *I*_*j*_ *=* 1, then *S*_*j*_∼*Bernoulli*(1 − *ψ*) ^+^ *Bernoulli*(*ϕ*) − *Bernoulli*(1 − *ψ*) × *Bernoulli*(*ϕ*), whereas if *I*_*j*_ *=* 0, then *S*_*j*_∼*Bernoulli*(*ϕ*). This yielded the crude proportion of asymptomatic individuals as

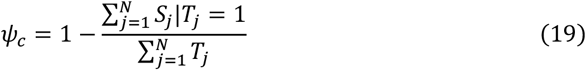

where the conditioning over *T*_*j*_ *=* 1 yields the number of symptomatic individuals among the test-positive subset (as is reported by surveys). Similarly, the crude proportion of individuals displaying symptoms among the test-negative subset is

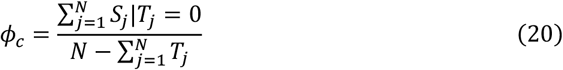

Together, *ρ*_*c*_, *ψ*_*c*_ and *ϕ*_*c*_, along with the distributions of *α* and *β*, which we constructed from reported means and confidence intervals of assay sensitivity and specificity (see Text S2), yielded the quantities reported by an individual survey that are needed to apply our formalism, namely, the quantities in the set *Q*. We repeated this multiple times, spanning ranges of the known quantities, to create diverse synthetic datasets reflective of the state of COVID-19 across reported serosurveys.

### Data from SARS-CoV-2 serosurveys

We searched Scopus, PubMed, and Google Scholar for serosurveys that provided data of all the quantities in *Q =* }*α, β, ρ*_*c*_, *ϕ*_*c*_, *ψ*_*c*_} required for applying equation (1). We obtained data from 50 serosurveys^18-20,22,28-73^ that met our criterion: We employed the terms ‘COVID-19,’ ‘SARS-CoV-2,’ ‘seroprevalence,’ ‘serological test,’ and ‘asymptomatic’ as search keywords when querying the databases. We chose retrospective cross-sectional serosurveys that determined the proportion of asymptomatic infections through interviews and questionnaires. We considered serosurveys in the early phase of the pandemic, before vaccination programs began, to eliminate any confounding effect of symptoms elicited by vaccines. Following previous considerations^99^, we restricted our analysis to studies with sample sizes ≥500 to minimize uncertainties in our calculations. We excluded studies on samples biased by symptom status, such as hospitalized patients, and focused on studies sampling the general population. All studies from where we extracted the data for analysis were published in peer-reviewed journals. We summarize the data in Table 1 and Table S1.

### Application of the formalism to data from serosurveys

In applying equation (1) to the datasets above, we accounted for the uncertainties in the quantities in *Q* and obtained estimates of *ψ* as well as its associated uncertainties. For the *i*^*th*^ dataset, let the reported quantities be *Q*_*i*_ *=* }*α*_*i*_, *β*_*i*_, *ρ*_*c*_, *ϕ*_*c*_, *ψ*_*c*_ }. Using these quantities and the reported sample size, *N*_*i*_, we produced 100000 virtual instantiations of the dataset by sampling the quantities from their associated distributions. Thus, for the *j*^*th*^ instantiation of the *i*^*th*^ dataset, we obtained the quantities 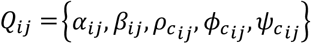 as:

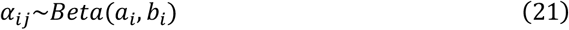

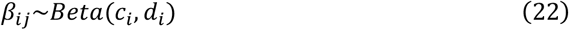

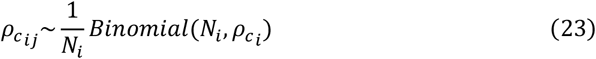

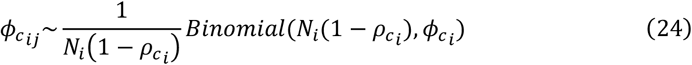

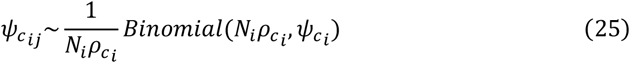

Here, the parameters *a*_*i*_ and *b*_*i*_ were chosen to recapitulate the known uncertainties in the sensitivity *α*_*i*_ (equation (21)) and the parameters *c*_*i*_ and *d*_*i*_ correspondingly for the specificity *β*_*i*_ (equation (22)). The identification of these uncertainties is described in Text S2, including for studies that used multiple assays^42,67^. The resulting parameter estimates are in Table S3. The seropositive fraction was assumed to follow a binomial distribution with parameters *N*_*i*_ and 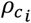 (equation (23)). Similarly, the symptomatic fraction among seronegative individuals followed a binomial with parameters 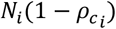, the sample size of seronegative individuals, and 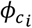 (equation (24)); and the asymptomatic fraction followed a binomial with parameters 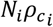, the sample size of seropositive individuals, and 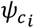 (equation (25)). Using the quantities sampled in the *j*^*th*^ instantiation in equation (1) yielded *ψ*_*ij*_, the estimate of the true asymptomatic proportion for the *j*^*th*^ instantiation. Repeating this 100000 times yielded the distribution of *ψ*_*i*_, the true proportion of asymptomatic infections associated with the *i*^*th*^ serosurvey, from which we obtained its median and 95% confidence intervals. (We employed 100000 repetitions as a conservative approach. In most cases, 1000 repetitions proved adequate.) Sampling may occasionally yield estimates of *ψ*_*ij*_ that are outside the range 0-1. Such estimates were neglected in computing the distribution of *ψ*_*i*_. We repeated this procedure for each of the 50 serosurvey datasets. We validated this procedure using synthetic datasets (Fig. 2).

### Extent of variation explained by symptom overlap

The distribution of *ψ*_*i*_ above also yielded the distribution of *η*_*i*_, the deviation of *ψ*_*i*_ from the crude estimate, 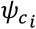, defined as 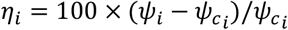. To determine the extent of this deviation explained by symptom overlap, we compared *η*_*i*_ with the prediction of the deviation due to symptom overlap alone (equation (2)): 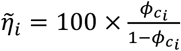. For this comparison, we considered an instantiation of 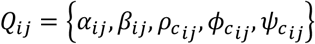 as described above and calculated *ψ*_*ij*_ using equation (1) and the corresponding 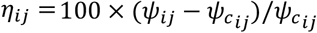. At the same time, we calculated *η*_*ij*_ estimated using equation (2) as 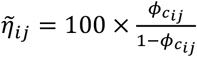. Repeating this for all *i* would yield an instantiation, the *j*^*th*^ instantiation, for all the 58 serosurveys. Comparing *η*_*ij*_ and 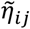 for all the *i*’s would yield an estimate of the extent to which the variation in *η*_*ij*_ is explained by symptom overlap in the *j*^*th*^ instantiation, quantified using *R*^*2*^. We repeated this procedure a 1000 times (letting *j =* 1,, *…*,1000) to obtain the associated distribution of *R*^*2*^ values. Note that the solid line in Fig. 4 is obtained by plotting the median 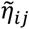 versus 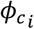 for all *i*.

### Data from PCR-based studies for validation

We first sought studies that conducted both PCR and serology tests, along with symptom assessment, on the same sample. At least a 2-week window for symptom assessment with the PCR test was necessary to identify presymptomatics.^74^ We restricted our search to studies before mass vaccination programs were initiated. From three comprehensive reviews on asymptomatic infections^9,17,100^, we identified 503 unique studies.^9,17,100^ Of these, 23 studies conducted antibody tests alongside PCR tests with >50 PCR-positive individuals, the latter we set as a cutoff for reliable statistics. None of the 23 studies, however, provided all the data (quantities in *Q*) required to estimate *ψ* using our formalism. We next conducted an independent search in the Scopus database for relevant studies using the following search key:

TITLE(“SARS-COV-2” OR “COVID-19”) AND TITLE-ABS-KEY(“PCR” OR “NUCLEIC ACID”) AND ALL(SYMPTOM*) AND TITLE-ABS-KEY(SERO* OR ANTIBODY) AND TITLE-ABS-KEY(PREVALENCE) AND PUBYEAR > 2019 AND PUBYEAR < 2023

From the resulting studies, we excluded reviews, studies determining accuracy of PCR or antibody tests, studies modeling *ψ* using estimates from other studies, studies involving non-human subjects, studies focused on non-SARS-CoV-2 infections, and studies not in English. We found 113 new studies that provided estimates of *ψ* using both PCR and antibody tests, of which 47 had test-positive individuals >50. Of the latter, three provided the necessary data to apply our formalism^78,101,102^. Upon further examination, we had to exclude two of these three studies: One study utilized PCR tests with a wide range of sensitivities (0.44 to 1), making its estimates unreliable.^101^ The second was restricted to a 2-week symptom recall period prior to antibody testing.^102^ Since antibody tests detect past infections, this short recall period risked misclassifying symptomatic individuals who had recovered earlier as asymptomatic. We employed the remaining study, on the 10 LTCFs in New Jersey^78^, for validation. The study had PCR/antigen test records performed ∼5 months prior to antibody testing. During the antibody testing, individuals were asked to recall symptoms developed over the past months. While *α* and *β* were reported for the antibody test, they were not available for the PCR test. We chose *α* for the PCR test based on a comprehensive systematic review which pooled PCR-based studies conducted until April 2020 and yielded a sensitivity of 0.87 (95% CI: 0.81–0.91).^103^ We set *β* to 1.00 (95% CI: 0.96– 1.00) based on multiple studies that suggested high specificity for PCR tests.^26,104^

Next, we looked for studies that reported PCR-based surveys along with a >2-week follow-up for symptom assessment conducted on samples representative of the general populations for which serosurvey data was available in our database (Table 1). We considered the same search methodology as above, drawing from the three systematic reviews^9,17,100^ and the Scopus database, and applied the same criteria above except for the requirement of both tests on the same sample. From the 503 studies from the systematic reviews and 113 from Scopus, three studies^79,105,106^ met our criteria. Of these studies, one considered symptoms only on the day of testing^105^ and another one week before testing^106^, potentially misclassifying presymptomatic individuals as asymptomatic, and were excluded. The remaining study, from Indiana, was selected.^79^ The study did not report values of *α* and *β*, which we therefore set to the pooled estimates above. Furthermore, the study did not report estimates of *ϕ*_*c*_. Recognizing that in the two-week follow-up period of the study, symptoms from an independent condition are unlikely, we simplified the derivation above, obviating the need for *ϕ*_*c*_. Specifically, when symptoms do not arise from other conditions, *P*[*S*^+^|*X*^−^] *=* 0 and *P*[*S*^−^|*X*^−^] *=* 1. This also implies that *P*[*S*^−^|*X*^+^] *= ψ*. Thus, equation (7) simplifies to 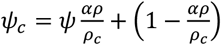, which upon combining with equation (4) yields 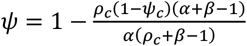. We employed the latter equation in place of equation (1) to obtain estimates of *ψ* for the Indiana study.

### Zika virus datasets

We collated data from 3 Zika virus serosurveys: The survey in Yap (2007)^80^ sampled 557 individuals, of which 414 tested positive for the infection. Of the latter, 258 declared no Zika virus symptoms. At the same time, 27 individuals from the test-negative group declared experiencing symptoms. The French Polynesia (2013-2014)^81^ survey sampled 196 individuals, of which 97 tested positive for the infection. Of the latter, 42 declared no symptoms, whereas of the test-negative group, 27 declared symptoms. Finally, the Puerto Rico (2016)^82^ study sampled 367 individuals, with 97 testing positive for the infection. Of the latter, 49 declared no symptoms. In contrast, 35 of the testnegative individuals declared experiencing symptoms. These observations yielded the inputs needed for the application of our formalism (Table 4). We compared the resulting estimates of *ψ* with those obtained by the Bayesian inference framework applied to the same datasets^7^ (Table 4).

### Web application development and benchmarking

We developed an easy-to-use, no-code web application, ShinyPsi, which enables the user to apply our formalism to their serosurvey data and download the results for future use. We used the Shiny^107,108^ framework in R to implement the application and then Shinylive^109^ for hosting it on a static webserver. The application can be accessed here along with instructions for its usage: https://therapeuticengglab.github.io/ShinyPsi/. The application carries out all the calculations within the web browser, eliminating the requirement of a dedicated Shiny server host. A major benefit, apart from ease of maintenance, is that no user data is collected. Thus, none of the epidemiological data that the user types in the application leaves their computers. Further, once the application page is loaded, the performance does not depend on internet speed or connectivity, as no output is returned from any remote server. The application runs effortlessly on most machines, yielding results in seconds. We tested ShinyPsi on standard laptops with MacOS (browser: Safari, 16 GB RAM), Windows OS (browser: Chrome, 16GB RAM) and Linux (browser: Firefox, 8GB RAM).

## Supporting information

All Supplementary Materials

## Data Availability

All data produced in the present work are contained in the manuscript

## ACKNOWLEDGMENTS

We thank Shreyas Joshi and Jeremie Guedj for comments.

## AUTHOR CONTRIBUTIONS

AT & NMD developed the formalism; AT collated the clinical datasets, performed the analysis, and wrote the first draft; BC developed the web application; SC, AJ & BC contributed to the analysis; NMD supervised the study and edited the draft.

## DATA AVAILABILITY

All data supporting the findings of this study are available within the paper and its Supplementary Information.

## CODE AVAILABILITY

We performed all calculations in R 4.3.3. The codes are available at: https://github.com/akshayytiwari/adjusted_psi

## REFERENCES

1. Shaikh, N., Swali, P. & Houben, R. Asymptomatic but infectious - The silent driver of pathogen transmission. A pragmatic review. Epidemics 44, 100704 (2023).

2. Montgomery, M.P., et al. The role of asymptomatic infections in influenza transmission: what do we really know. Lancet Infect. Dis. 24, e394–e404 (2024).

3. Glynn, J.R., et al. Asymptomatic infection and unrecognised Ebola virus disease in Ebola-affected households in Sierra Leone: a cross-sectional study using a new non-invasive assay for antibodies to Ebola virus. Lancet Infect. Dis. 17, 645–653 (2017).

4. Chen, I., et al. “Asymptomatic” malaria: A chronic and debilitating infection that should be treated. PLoS Med. 13, e1001942 (2016).

5. Leung, N.H.L., Xu, C., Ip, D.K.M. & Cowling, B.J. Review article: The fraction of influenza virus infections that are asymptomatic: A Systematic review and meta-analysis. Epidemiology 26, 862–872 (2015).

6. Wang, T.E., Lin, C.Y., King, C.C. & Lee, W.C. Estimating pathogen-specific asymptomatic ratios. Epidemiology 21, 726–728 (2010).

7. Mitchell, P.K., et al. Reassessing serosurvey-based estimates of the symptomatic proportion of Zika virus infections. Amer. J. Epidemiol. 188, 206–213 (2018).

8. Johansson, M.A., et al. SARS-CoV-2 transmission from people without COVID-19 symptoms. JAMA Netw. Open 4, e2035057 (2021).

9. Buitrago-Garcia, D., et al. Occurrence and transmission potential of asymptomatic and presymptomatic SARS-CoV-2 infections: Update of a living systematic review and meta-analysis. PLoS Med 19, e1003987 (2022).

10. Cramer, E.Y., et al. Evaluation of individual and ensemble probabilistic forecasts of COVID-19 mortality in the United States. Proc. Natl. Acad. Sci. USA 119, e2113561119 (2022).

11. Ganser, I., Buckeridge, D.L., Heffernan, J., Prague, M. & Thiébaut, R. Estimating the population effectiveness of interventions against COVID-19 in France: A modelling study. Epidemics 46, 100744 (2024).

12. Buitrago-Garcia, D., et al. Occurrence and transmission potential of asymptomatic and presymptomatic SARS-CoV-2 infections: A living systematic review and meta-analysis. PLoS Med. 17, e1003346 (2020).

13. Bertozzi, A.L., Franco, E., Mohler, G., Short, M.B. & Sledge, D. The challenges of modeling and forecasting the spread of COVID-19. Proc. Natl. Acad. Sci. USA 117, 16732–16738 (2020).

14. Loo, S.L., et al. The US COVID-19 and Influenza Scenario Modeling Hubs: Delivering long-term projections to guide policy. Epidemics 46, 100738 (2024).

15. Kissler, S.M., Tedijanto, C., Goldstein, E., Grad, Y.H. & Lipsitch, M. Projecting the transmission dynamics of SARS-CoV-2 through the postpandemic period. Science 368, 860–868 (2020).

16. Oran, D.P. & Topol, E.J. The proportion of SARS-CoV-2 infections that are asymptomatic : A systematic review. Ann. Intern. Med. 174, 655–662 (2021).

17. Sah, P., et al. Asymptomatic SARS-CoV-2 infection: A systematic review and meta-analysis. Proc. Natl. Acad. Sci. USA 118, e2109229118 (2021).

18. Vos, E.R.A., et al. Nationwide seroprevalence of SARS-CoV-2 and identification of risk factors in the general population of the Netherlands during the first epidemic wave. J Epidemiol. Community Health 75, 489–495 (2020).

19. Sullivan, P.S., et al. Severe acute respiratory syndrome coronavirus 2 cumulative incidence, United States, August 2020-December 2020. Clin. Infect. Dis. 74, 1141–1150 (2022).

20. Chamberlain, A.T., et al. Cumulative incidence of SARS-CoV-2 infections among adults in Georgia, United States, August to December 2020. J. Infect. Dis. 225, 396–403 (2022).

21. Rogan, W.J. & Gladen, B. Estimating prevalence from the results of a screening test. Amer. J. Epidemiol. 107, 71–76 (1978).

22. Canto e Castro, L., et al. Prevalence of SARS-CoV-2 antibodies after first 6 months of COVID-19 pandemic, Portugal. Emerg. Infect. Dis. 27, 2878 (2021).

23. McDonald, S.A., et al. Estimating the asymptomatic proportion of SARS-CoV-2 infection in the general population: Analysis of nationwide serosurvey data in the Netherlands. Eur. J. Epidemiol 36, 735–739 (2021).

24. Arora, R.K., et al. SeroTracker: a global SARS-CoV-2 seroprevalence dashboard. Lancet Infect. Dis. 21, e75–e76 (2021).

25. Kucirka, L., Lauer, S., Laeyendecker, O., Boon, D. & Lessler, J. Variation in false-negative rate of reverse transcriptase polymerase chain reaction–based SARS-CoV-2 tests by time since exposure. Ann. Intern. Med. 173, 262–267 (2020).

26. Mair, M.D., et al. A systematic review and meta-analysis comparing the diagnostic accuracy of initial RT-PCR and CT scan in suspected COVID-19 patients. Br. J. Radiol. 94, 20201039 (2021).

27. Lin, L.I.K. A concordance correlation coefficient to evaluate reproducibility. Biometrics 45, 255–268 (1989).

28. Menezes, A.M.B., et al. High prevalence of symptoms among Brazilian subjects with antibodies against SARS-CoV-2. Sci. Rep. 11, 13279 (2021).

29. Silva, A.A.M.D., et al. Population-based seroprevalence of SARS-CoV-2 and the herd immunity threshold in Maranhao. Rev. Saude Publica 54, 131 (2020).

30. Terças-Trettel, A.C.P., Muraro, A.P., Andrade, A.C.S. & Oliveira, E.C. Self-reported symptoms and seroprevalence against SARS-CoV-2 in the population of Mato Grosso: a household-based survey in 2020. Rev. Assoc. Med. Bras. (1992) 68, 928–934 (2022).

31. Albuquerque, J.O.M., et al. Prevalence evolution of SARS-CoV-2 infection in the city of Sao Paulo, 2020-2021. Rev. Saude Publica 55, 62 (2021).

32. Nwosu, K., et al. SARS-CoV-2 antibody seroprevalence and associated risk factors in an urban district in Cameroon. Nat. Commun. 12, 5851 (2021).

33. Vial, P., et al. Seroprevalence, spatial distribution, and social determinants of SARS-CoV-2 in three urban centers of Chile. BMC Infect. Dis. 22, 99 (2022).

34. Li, Z., et al. Antibody seroprevalence in the epicenter Wuhan, Hubei, and six selected provinces after containment of the first epidemic wave of COVID-19 in China. Lancet Reg. Health. West. Pac. 8, 100094 (2021).

35. Garay, E., et al. SARS-CoV-2 in eight municipalities of the Colombian tropics: high immunity, clinical and sociodemographic outcomes. Trans. R. Soc. Trop. Med. Hyg. 116, 139–147 (2022).

36. Serrano-Coll, H., et al. High prevalence of SARS-CoV-2 in an Indigenous community of the Colombian Amazon region. Trop. Med. Infect. Dis. 6, 191 (2021).

37. Espenhain, L., et al. Prevalence of SARS-CoV-2 antibodies in Denmark: Nationwide, population-based seroepidemiological study. Eur. J. Epidemiol. 36, 715–725 (2021).

38. Carrat, F., et al. Antibody status and cumulative incidence of SARS-CoV-2 infection among adults in three regions of France following the first lockdown and associated risk factors: a multicohort study. Int. J. Epidemiol. 50, 1458–1472 (2021).

39. Rouquette, A., et al. Comparison of depression and anxiety following self-reported COVID-19-like symptoms vs SARS-CoV-2 seropositivity in France. JAMA Netw. Open 6, e2312892 (2023).

40. Beaumont, A., et al. Seroprevalence of anti-SARS-CoV-2 antibodies after the first wave of the COVID-19 pandemic in a vulnerable population in France: A cross-sectional study. BMJ Open 11, e053201 (2021).

41. Santos-Hövener, C., et al. Serology- and PCR-based cumulative incidence of SARS-CoV-2 infection in adults in a successfully contained early hotspot (CoMoLo study), Germany, May to June 2020. Euro. Surveill. 25, 2001752 (2020).

42. Weis, S., et al. Antibody response using six different serological assays in a completely PCR-tested community after a coronavirus disease 2019 outbreak-the CoNAN study. Clin. Microbiol. Infect. 27, 470 e471–470 e479 (2021).

43. Merkely, B., et al. Novel coronavirus epidemic in the Hungarian population, a cross-sectional nationwide survey to support the exit policy in Hungary. Geroscience 42, 1063–1074 (2020).

44. Murhekar, M.V., et al. SARS-CoV-2 antibody seroprevalence in India, August-September, 2020: Findings from the second nationwide household serosurvey. Lancet Glob. Health 9, e257–e266 (2021).

45. Selvaraju, S., et al. Population-based serosurvey for severe acute respiratory syndrome coronavirus 2 transmission, Chennai, India. Emerg. Infect. Dis. 27, 586–589 (2021).

46. Kumar, M.S., et al. Monitoring the trend of SARS-CoV-2 seroprevalence in Chennai, India, July and October 2020. Trans. R. Soc. Trop. Med. Hyg. 115, 1350–1352 (2021).

47. Sharma, N., et al. The seroprevalence of severe acute respiratory syndrome coronavirus 2 in Delhi, India: a repeated population-based seroepidemiological study. Trans. R. Soc. Trop. Med. Hyg. 116, 242–251 (2022).

48. Kumar, D., et al. Seroprevalence of anti SARS-CoV-2 IgG antibodies among adults in Jammu district, India: A community-based study. Indian J. Med. Res. 155, 171–177 (2022).

49. Khan, S.M.S., et al. Seroprevalence of SARS-CoV-2-specific IgG antibodies in Kashmir, India, 7 months after the first reported local COVID-19 case: results of a population-based seroprevalence survey from October to November 2020. BMJ Open 11, e053791 (2021).

50. Poustchi, H., et al. SARS-CoV-2 antibody seroprevalence in the general population and high-risk occupational groups across 18 cities in Iran: a population-based cross-sectional study. Lancet Infect. Dis. 21, 473–481 (2021).

51. Shadmani, F.K., et al. Seroprevalence of SARS-Cov-2 virus infection in Kermanshah, Iran: a population-based cross-sectional study. Open Public Health J. 16, e187494452302011 (2023).

52. Heavey, L., et al. The study to investigate COVID-19 infection in people living in Ireland (SCOPI): A seroprevalence study, June to July 2020. Euro Surveill. 26, 2001741 (2021).

53. Melotti, R., et al. Prevalence and determinants of serum antibodies to SARS-CoV-2 in the general population of the Gardena valley. Epidemiol. Infect. 149, e194 (2021).

54. Pagani, G., et al. Prevalence of SARS-CoV-2 in an area of unrestricted viral circulation: Mass seroepidemiological screening in Castiglione d’Adda, Italy. PLoS One 16, e0246513 (2021).

55. Ngere, I., et al. High seroprevalence of SARS-CoV-2 but low infection fatality ratio eight months after introduction in Nairobi, Kenya. Int. J. Infect. Dis. 112, 25–34 (2021).

56. Abdul-Raheem, R., et al. A sero-epidemiological study after two waves of the COVID-19 epidemic. Asian Pac. J. Allergy Immunol. 42, 270–275 (2021).

57. Sagara, I., et al. Rapidly increasing severe acute respiratory syndrome coronavirus 2 seroprevalence and limited clinical disease in 3 Malian communities: A prospective cohort study. Clin. Infect. Dis. 74, 1030–1038 (2022).

58. Basto-Abreu, A., et al. Nationally representative SARS-CoV-2 antibody prevalence estimates after the first epidemic wave in Mexico. Nat. Commun. 13, 589 (2022).

59. Arnaldo, P., et al. Prevalence of severe acute respiratory syndrome coronavirus 2 (SARS-CoV-2) antibodies in the Mozambican population: A cross-sectional serologic study in 3 cities, July-August 2020. Clin. Infect. Dis. 75, S285–S293 (2022).

60. Okpala, O.V., et al. Population seroprevalence of SARS-CoV-2 antibodies in Anambra State, South-East, Nigeria. Int. J. Infect. Dis. 110, 171–178 (2021).

61. Nisar, M.I., et al. Serial population-based serosurveys for COVID-19 in two neighbourhoods of Karachi, Pakistan. Int. J. Infect. Dis. 106, 176–182 (2021).

62. Huamaní, C., Velásquez, L., Montes, S., Mayanga-Herrera, A. & Bernabé-Ortiz, A. SARS-CoV-2 seroprevalence in a high-altitude setting in Peru: adult population-based cross-sectional study. PeerJ 9, e12149 (2021).

63. Díaz-Vélez, C., et al. SARS-CoV-2 seroprevalence study in Lambayeque, Peru. June-July 2020. PeerJ 9, e11210 (2021).

64. Reyes-Vega, M.F., et al. SARS-CoV-2 prevalence associated to low socioeconomic status and overcrowding in an LMIC megacity: A population-based seroepidemiological survey in Lima, Peru. EClinMed 34, 100801 (2021).

65. Moyano, L.M., et al. SARS-CoV-2 seroprevalence on the north coast of Peru: A cross-sectional study after the first wave. PLoS Negl. Trop. Dis. 17, e0010794 (2023).

66. Kislaya, I., et al. Seroprevalence of SARS-CoV-2 Infection in Portugal in May-July 2020: Results of the First National Serological Survey (ISNCOVID-19). Acta Med. Port. 34, 87–94 (2021).

67. Talla, C., et al. Seroprevalence of anti-SARS-CoV-2 antibodies in Senegal: a national population-based cross-sectional survey, between October and November 2020. IJID Reg 3, 117–125 (2022).

68. Pérez-Gómez, B., et al. ENE-COVID nationwide serosurvey served to characterize asymptomatic infections and to develop a symptom-based risk score to predict COVID-19. J. Clin. Epidemiol. 139, 240–254 (2021).

69. Richard, A., et al. Seroprevalence of anti-SARS-CoV-2 IgG antibodies, risk factors for infection and associated symptoms in Geneva, Switzerland: a population-based study. Scand. J. Public Health 50, 124–135 (2022).

70. Alsuwaidi, A.R., et al. Seroprevalence of COVID-19 infection in the Emirate of Abu Dhabi, United Arab Emirates: a population-based cross-sectional study. Int. J. Epidemiol. 50, 1077–1090 (2021).

71. Ward, H., et al. Prevalence of antibody positivity to SARS-CoV-2 following the first peak of infection in England: Serial cross-sectional studies of 365,000 adults. Lancet Reg. Health Eur. 4, 100098 (2021).

72. Lamba, K., et al. SARS-CoV-2 cumulative incidence and period seroprevalence: Results from a statewide population-based serosurvey in California. Open Forum Infect. Dis. 8, ofab379 (2021).

73. Pathela, P., et al. Seroprevalence of severe acute respiratory syndrome coronavirus 2 following the largest initial epidemic wave in the United States: Findings from New York City, 13 May to 21 July 2020. J. Infect. Dis. 224, 196–206 (2021).

74. Meyerowitz, E.A., Richterman, A., Bogoch, II, Low, N. & Cevik, M. Towards an accurate and systematic characterisation of persistently asymptomatic infection with SARS-CoV-2. Lancet Infect. Dis. 21, e163–e169 (2021).

75. Dan, J.M., et al. Immunological memory to SARS-CoV-2 assessed for up to 8 months after infection. Science 371, eabf4063 (2021).

76. Davies, N.G., et al. Age-dependent effects in the transmission and control of COVID-19 epidemics. Nat. Med. 26, 1205–1211 (2020).

77. Poletti, P., et al. Association of age with likelihood of developing symptoms and critical disease among close contacts exposed to patients with confirmed SARS-CoV-2 infection in Italy. JAMA Netw. Open 4, e211085 (2021).

78. Friedman, S.M., et al. Antibody seroprevalence, infection and surveillance for SARS-CoV-2 in residents and staff of New Jersey long-term care facilities. J. Community Health 47, 774–782 (2022).

79. Menachemi, N., et al. Population point prevalence of SARS-CoV-2 infection based on a statewide random sample - Indiana, April 25-29, 2020. Morb. Mortal. Wkly. Rep. 69, 960–964 (2020).

80. Duffy, M.R., et al. Zika virus outbreak on Yap Island, Federated States of Micronesia. New Engl. J. Med. 360, 2536–2543 (2009).

81. Aubry, M., et al. Zika virus seroprevalence, French Polynesia, 2014–2015. Emerging Infect. Dis. 23, 669 (2017).

82. Lozier, M.J., et al. Differences in prevalence of symptomatic Zika virus infection, by age and sex—Puerto Rico, 2016. J. Infect. Dis. 217, 1678–1689 (2018).

83. Augusto, D.G., et al. A common allele of HLA is associated with asymptomatic SARS-CoV-2 infection. Nature 620, 128–136 (2023).

84. Marchal, A., et al. Lack of association between classical HLA genes and asymptomatic SARS-CoV-2 infection. HGG Adv. 5, 100300 (2024).

85. Chowdhury, S., Tiwari, A., James, A., Chatterjee, B. & Dixit, N.M. Asymptomatic SARS-CoV-2 infections tend to occur less frequently in developed nations. medRxiv, 2023.2012.2014.23299954 (2023).

86. Owens, K., Esmaeili, S. & Schiffer, J.T. Heterogeneous SARS-CoV-2 kinetics due to variable timing and intensity of immune responses. JCI Insight 9, e176286 (2024).

87. Chatterjee, B., Singh Sandhu, H. & Dixit, N.M. Modeling recapitulates the heterogeneous outcomes of SARS-CoV-2 infection and quantifies the differences in the innate immune and CD8 T-cell responses between patients experiencing mild and severe symptoms. PLoS Pathog. 18, e1010630 (2022).

88. Zhu, C., Pang, S., Liu, J. & Duan, Q. Current progress, challenges and prospects in the development of COVID-19 vaccines. Drugs 84, 403–423 (2024).

89. Khoury, D.S., et al. Neutralizing antibody levels are highly predictive of immune protection from symptomatic SARS-CoV-2 infection. Nat. Med. 27, 1205–1211 (2021).

90. Khoury, D.S., et al. Predicting the efficacy of variant-modified COVID-19 vaccine boosters. Nat. Med. 29, 574–578 (2023).

91. Padmanabhan, P., Desikan, R. & Dixit, N.M. Modeling how antibody responses may determine the efficacy of COVID-19 vaccines. Nat. Comput. Sci. 2, 123–131 (2022).

92. Lumley, S.F., et al. Antibody status and incidence of SARS-CoV-2 infection in health care workers. N. Engl. J. Med. 384, 533–540 (2021).

93. Abu-Raddad, L.J., et al. SARS-CoV-2 antibody-positivity protects against reinfection for at least seven months with 95% efficacy. EClinMed 35, 100861 (2021).

94. Pawlowski, C., et al. SARS-CoV-2 and influenza coinfection throughout the COVID-19 pandemic: an assessment of coinfection rates, cohort characteristics, and clinical outcomes. PNAS Nexus 1, pgac071 (2022).

95. Olsen, S.J., et al. Changes in influenza and other respiratory virus activity during the COVID-19 pandemic—United States, 2020–2021. Amer. J. Transplant. 21, 3481–3486 (2021).

96. Almadhoon, H.W., et al. The effect of influenza vaccine in reducing the severity of clinical outcomes in patients with COVID-19: a systematic review and meta-analysis. Sci. Rep. 12, 14266 (2022).

97. Korosec, C.S., et al. Long-term durability of immune responses to the BNT162b2 and mRNA-1273 vaccines based on dosage, age and sex. Sci. Rep. 12, 21232 (2022).

98. Peluso, M.J., et al. SARS-CoV-2 antibody magnitude and detectability are driven by disease severity, timing, and assay. Sci Adv 7, eabh3409 (2021).

99. Ioannidis, J.P.A. Infection fatality rate of COVID-19 inferred from seroprevalence data. Bull. World Health Organ. 99, 19–33F (2021).

100. Ma, Q., et al. Global percentage of asymptomatic SARS-CoV-2 infections among the tested population and individuals with confirmed COVID-19 diagnosis: a systematic review and meta-analysis. JAMA Netw. Open 4, e2137257 (2021).

101. Garralda Fernandez, J., et al. Impact of SARS-CoV-2 pandemic among health care workers in a secondary teaching hospital in Spain. PLoS One 16, e0245001 (2021).

102. Al-Thani, M.H., et al. SARS-CoV-2 Infection Is at Herd Immunity in the Majority Segment of the Population of Qatar. Open Forum Infect. Dis. 8, ofab221 (2021).

103. Arevalo-Rodriguez, I., et al. False-negative results of initial RT-PCR assays for COVID-19: A systematic review. PLoS ONE 15, e0242958 (2020).

104. Skittrall, J.P., et al. Specificity and positive predictive value of SARS-CoV-2 nucleic acid amplification testing in a low-prevalence setting. Clin. Microbiol. Infect. 27, 469.e469-469.e415 (2021).

105. Petersen, I. & Phillips, A. Three quarters of people with SARS-CoV-2 infection are asymptomatic: analysis of English household survey data. Clin. Epidemiol. 12, 1039–1043 (2020).

106. Riley, S., et al. Resurgence of SARS-CoV-2: Detection by community viral surveillance. Science 372, 990–995 (2021).

107. Chang, W., et al. Shiny: web application framework for R. https://shiny.rstudio.com/ (2021).

108. Wickham, H. Mastering Shiny, (O’Reilly Media, Inc., 2021).

109. Schloerke, B., Chang, W., Stagg, G. & Aden-Buie, G. Shinylive: Run’shiny’applications in the browser. https://posit-dev.github.io/r-shinylive/ (2023).

