## Supplementary material for "A fast and accurate method for estimating the proportion of asymptomatic infections from serosurveys: Application to SARS-CoV-2 and beyond": All Supplementary Materials

This document contains:

Text notes S1-S2;

Tables S1-S3;

Figures S1-S7;

References: 14

**Text S1. Effect of time-varying assay characteristics on estimates of proportion of asymptomatic infections**

With nucleic acid-based testing, the sensitivity, $\alpha$, can vary substantially from the time of exposure, due to associated changes in viral load and other biological or technical factors.[1] A study analyzing data from seven earlier studies of RT-PCR samples (n=1330) estimated that $\alpha$ varied from 33% on the day before symptom onset (typically 4 days after exposure) to a maximum of ~80% three days after symptom onset and declined again to ~34% two-three weeks later.[1] The same study found that the specificity, $\beta$, displayed little variation over the same duration, and remained around 90-95%, consistent with independent studies that found $\beta$ for SARS-CoV-2 to be high (median $\beta$=1; 95% CI: (0.96, 1.00))[2]. Here, we examined how surveys conducted at different time points from exposure may lead to errors in the estimation of $\psi$ due to the associated variation in assay characteristics. We generated synthetic data reflecting surveys conducted at different time points, following the procedure outlined in Methods. We set $\psi$ (or $\psi_{true}$) and $\rho$ $(\text{or } \rho_{true})$ to their median values from the 50 serosurveys we studied (Table 1). Given the short duration (<3 weeks) of PCR-based surveys, we assumed that confounding from overlapping symptoms may be negligible. We thus let $\phi = 0$. As a further simplification, we set $\beta = 1$ following the above estimates. $\alpha$ was chosen based on the measurements above. We then estimated $\psi$ using our formalism and calculated the error from the true value, $\frac{\psi - \psi_{true}}{\psi_{true}} \times 100$. We found that the error was lowest (6.5%, IQR: (5.2%–7.7%)) around day 8, when $\alpha$ was the highest (0.80, 95% CI: (0.70, 0.89)), and increased at earlier (12.4%, IQR: (10.4%, 14.1%) and later stages (13.9%, IQR: (11.9%, 15.7%)) of infection when $\alpha$ was low (~0.33) (Fig. S1). These results indicate that the estimate of $\psi$ is most accurate when

$\alpha$ is near its peak. These trends may inform PCR-based survey designs and help better interpret their findings. Note that antibodies last much longer (months)[3], resulting in much less of a variation in $\alpha$ and $\beta$ with time from infection compared to PCR-based tests.

**Text S2. Distributions of sensitivity and specificity of test assays**

Here, we describe how we estimated the distributions of sensitivity, $\alpha$, and specificity, $\beta$, of the test assays used in the 50 serosurveys we studied (Table 1). Recall that the surveys yielded 58 estimates of $\psi_c$ as four studies reported estimates at three different time points. We identified the assays used for each of these estimates and collated reported estimates and uncertainties of $\alpha$ and $\beta$. We found that in 50 instances (not all overlapping), mean values and 95% confidence intervals were available for $\alpha$ and $\beta$. Based on these values, we identified parameters–$a$, $b, c$ and $d$–of the Beta distributions for $\alpha \sim Beta(a, b)$ and $\beta \sim Beta(c, d)$ that matched the corresponding means and confidence intervals.

In one instance each for $\alpha$[4] and $\beta$[5] and 5 instances for both[6-10], mean values were available but not confidence intervals. In such instances, we chose the parameters of the distributions, $Beta(\mu\kappa, (1-\mu)\kappa)$, to yield the mean $\mu$ and set $\kappa = 20$ (typically chosen to lie between 10 and 50[11]) for a sufficiently wide confidence interval, as a conservative estimate. (Varying $\kappa$ did not affect our estimates.)

In two instances, Weis et al.[12] and Talla et al.[13], multiple assays were used to detect seropositivity. We considered these cases separately and derived expressions to estimate the corresponding distributions of sensitivity and specificity, which we describe below.

The resulting distributions of $\alpha$ and $\beta$ for all the 58 instances are in Table S3. We employed these distributions for estimating uncertainties in $\psi$ (Methods).

***Weis et al.*[12]**

Weis et al. used six different assays on each survey participant and categorized the participant as seropositive if at least two assays yielded a positive result. For each assay, both sensitivity and specificity with their respective 95% confidence intervals were reported. Here, we developed a way to estimate the overall sensitivity and specificity of the testing method.

For an individual participant, let $R(0 \leq R \leq 6)$ denote the number of assays that yield a positive result. Let $X^+$ indicate infection due to the pathogen of interest, $X$. The overall sensitivity, $\alpha_o$ is the probability of $R \geq 2$, conditional on $X^+$. Thus,

$$\alpha_o = P(R \geq 2|X^+) = 1 - P(R = 0|X^+) - P(R = 1|X^+) \qquad (S1)$$

If $\alpha_k$ were the sensitivity of the $k^{th}$ $(1 \leq k \leq 6)$ assay, then the probability of no assay yielding a positive result given the infection would be

$$P(R = 0|X^+) = \prod_{k=1}^{6}(1 - \alpha_k) \qquad (S2)$$

Similarly, the probability of exactly one positive result of the six tests given the infection would be

$$P(R = 1|X^+) = \sum_{k=1}^{6} \alpha_k \prod_{j \neq k}^{6}(1 - \alpha_j) \qquad (S3)$$

Combining the three equations above yields

$$\alpha_o = 1 - \prod_{k=1}^{6}(1 - \alpha_k) - \sum_{k=1}^{6} \alpha_k \prod_{j \neq k}^{6}(1 - \alpha_j) \qquad (S4)$$

Equation (S4) yielded an expression for the overall sensitivity of the method, $\alpha_o$, given the sensitivities of the individual assays. To obtain the distribution of $\alpha_o$, we performed Monte Carlo simulations sampling the individual sensitivities, $\alpha_k$, from their respective distributions and estimating $\alpha_o$ using equation (S4). The distributions of the individual assays are listed in Table S3. This yielded the median $\alpha_o = 1$ and the 95% confidence interval of $(1,1)$. Note that the probability of at least two of the six tests yielding a positive result following an infection is very high, explaining the high sensitivity of the method and justifying the use of multiple assays.

Similarly, we estimated the overall specificity, $\beta_o$, given the specificities of the individual assays as follows. Seronegativity in the method was defined to occur when $R < 2$, conditional on $X^-$, the absence of infection by $X$. Thus,

$$\beta_o = P(R < 2|X^-) = P(R = 0|X^-) + P(R = 1|X^-) \qquad (S5)$$

If $\beta_k$ were the specificity of the $k^{th}$ $(1 \leq k \leq 6)$ assay, then the probability of no assay yielding a positive result given the absence of infection would be

$$P(R = 0|X^-) = \prod_{k=1}^{6} \beta_k \qquad (S6)$$

Similarly, the probability of exactly one positive result given the absence of infection would be

$$P(R = 1|X^-) = \sum_{k=1}^{6} (1 - \beta_k) \prod_{j \neq k}^{6} \beta_j \qquad (S7)$$

Combining the three latter equations yields,

$$\beta_o = \prod_{k=1}^{6} \beta_k + \sum_{k=1}^{6} (1 - \beta_k) \prod_{j \neq k}^{6} \beta_j \qquad (S8)$$

Equation (S8) yielded an expression for the overall specificity of the method, $\beta_o$, given the specificities of the individual assays. To obtain the distribution of $\beta_o$, we again performed Monte Carlo simulations sampling the individual specificities, $\beta_k$, from their respective distributions (Table S3) and estimating $\beta_o$ using equation (S8). This yielded the median $\beta_o = 0.999$ and the 95% confidence interval of $(0.997, 1.000)$, the high specificity reiterating the advantage of using multiple assays.

### *Talla et al.*[13]

Talla et al. subjected individual participants to two assays to detect the infection. They provided explicit formulae for the overall sensitivity and specificity of their dual-assay detection method:

$$\alpha_o = \alpha_1 \alpha_2 \qquad (S9)$$

$$\beta_o = \beta_1 + \beta_2 - \beta_1 \beta_2 \qquad (S10)$$

where subscripts 1 and 2 refer to test 1 (OMEGA COVID-19 IgG) and test 2 (IDVet assay), respectively. The mean values and 95% confidence intervals of the sensitivity and specificity of both assays were reported. We thus constructed Beta distributions for each of these quantities and, following the procedure above, performed Monte Carlo simulations by sampling from these distributions and applying equations (S9) and (S10) to obtain distributions of $\alpha_o$ and $\beta_o$. This yielded the median $\alpha_o = 0.805$ and its 95% confidence interval to be $(0.618,\ 0.926)$, and the median $\beta_o = 0.986$ with the 95% confidence interval of $(0.922,\ 0.999)$.

**Table S1. Serosurvey details.** Descriptions of the locations involved in each serosurvey, their geographical scope, the median age of participants, and the symptom recall periods employed are listed. N: nation-wide; S: subnational; L: local. See Table 1 for other details and study citations.

| **Study** | **Scope** | **Area(s)** | **Median age (years)** | **Recall period (weeks)** |
|---|---|---|---|---|
| Menezes et al. | N | Sentinel cities in 26 Brazilian states and the Federal District. | 43.3* | 12 |
| Silva et al. | S | The state of Maranhão, including the capital São Luís and a range of small to large municipalities. | 44.4* | 26 |
| Terças-Trettel et al. | S | Mato Grosso, a Midwestern Brazilian state, covering pole municipalities of the socioeconomic regions of Mato Grosso and the main cities. | 46.2 | 31 |
| Albuquerque et al. | L | City of São Paulo. | 47.3* | 28† |
| Albuquerque et al. | L | City of São Paulo. | 47.1* | 28† |
| Albuquerque et al. | L | City of São Paulo. | 49.9* | 28† |
| Nwosu et al. | L | Cité Verte, a health district of Yaoundé, Cameroon. | 26.0 | 36 |
| Vial et al. | L | Three urban areas in central Chile: Greater Santiago, Coquimbo-La Serena, and Talca. | 41.6 | 39† |
| Li et al. | L | China across Wuhan, the rest of Hubei province, and six other major provinces or municipalities. | 41.9* | 19 |
| Garay et al. | L | The department of Córdoba, Colombia, specifically targeting its eight most populous municipalities. | 42.0 | 13 |
| Serrano-Coll et al. | N | Mitú, a municipality in the Colombian Amazon near the border with Brazil. | 35.5* | 46† |
| Espenhain et al | S | Denmark, as part of a nationwide seroprevalence survey using the national civil registry. | 51.6* | 36 |
| Carrat et al. | L | Three French regions: Île-de-France, Grand Est, and Nouvelle-Aquitaine. | 57.1* | 2 |

| | | | | |
|---|---|---|---|---|
| Rouquette et al. | N | France using a nationally representative sample drawn from the administrative and tax database. | 51.1 | 28 |
| Beaumont et al. | S | Three neighbourhoods of Perpignan, a city in southern France. | 42.4* | 19 |
| Santos-Hövener et al. | N | Kupferzell, a small town in southern Germany. | 45.9* | 17 |
| Weis et al. | L | Neustadt am Rennsteig, a village in Thuringia, central Germany. | 60.0 | 6 |
| Merkely et al. | L | Hungary, using a nationally representative sample covering all regions and settlement types. | 48.7 | 18 |
| Murhekar et al. | L | Seventy districts in India, covering 700 villages or urban wards as part of a national serosurvey. | 37.0 | 27 |
| Selvaraju et al. | N | Greater Chennai Corporation, a major urban area in southern India. | 41.1 | 13 |
| Kumar et al. | N | Fifty-one selected wards of Chennai, a major metropolitan city in southern India. | 39.8 | 37† |
| Sharma et al. | L | Delhi, covering its 11 districts and 280 wards. | 35.0* | 27† |
| Sharma et al. | L | Delhi, covering its 11 districts and 280 wards. | 35.0* | 35† |
| Sharma et al. | L | Delhi, covering its 11 districts and 280 wards. | 34.6* | 31† |
| Kumar et al. | L | Ten health blocks of Jammu district, India. | 40.2 | 26 |
| Khan et al. | S | All 10 districts of the Kashmir Valley in northern India. | 42.0* | 13 |
| Poustchi et al. | S | Eighteen cities in 17 provinces of Iran, to estimate nationwide SARS-CoV-2 seroprevalence. | — | 12 |
| Shadmani et al. | S | Kermanshah province, western Iran, covering 14 cities within the province. | 35.7 | 44† |
| Heavey et al. | L | Two Irish counties, Dublin and Sligo, representing areas of high and low COVID-19 incidence. | 43.3* | 18 |
| Melotti et al. | S | Three municipalities of the Gardena Valley, a winter tourist region in northern Italy. | 45.1 | 18 |

| | | | | |
|---|---|---|---|---|
| Pagani et al. | N | Castiglione d'Adda, a municipality in Lombardy, northern Italy. | 48.0 | 17 |
| Ngere et al. | S | Nairobi City County, Kenya, covering all 17 of its administrative sub-counties. | 26.0 | 52 |
| Abdul-Raheem et al. | L | The Greater Malé Area, the urban capital region of the Maldives. | 27.6* | 39† |
| Sagara et al. | L | Three communities in Mali: Sotuba, Bancoumana, and Donéguébougou, representing urban and rural settings. | 14.0 | 27 |
| Basto-Abreu et al. | L | Mexico, as part of the nationally representative 2020 health and nutrition survey. | 39.2* | 30 |
| Arnaldo et al. | L | Pemba city of Cabo Delgado province. | 21.0 | 4 |
| Arnaldo et al. | L | Maputo city in the province of Maputo City. | 28.0 | 4 |
| Arnaldo et al. | L | Quelimane city of Zambézia province. | 21.0 | 4 |
| Vos et al. | L | The Netherlands, using a nationally representative sample from 40 municipalities in five regions. | 42.1* | 12† |
| Okpala et al. | L | All local government areas of Anambra state in southeastern Nigeria. | 30.2* | 45† |
| Nisar et al. | N | Selected union councils of District East and District Malir in Karachi, Pakistan. | 26.3 | 9 |
| Huamani et al. | L | The Cusco region of southeastern Peru, including Cusco City, nearby districts (Santiago, San Jerónimo, San Sebastián, and Wanchaq), and Quillabamba. | 42.5 | 13 |
| Díaz-Vélez et al. | L | The Lambayeque region of northern Peru, covering 38 districts. | 42.8* | 24† |
| Reyes-Vega et al. | L | The Lima metropolitan area of Peru, covering 43 districts of Lima and 7 districts of Callao provinces. | 37.3* | 22† |
| Moyano et al. | N | Puerto Pizarro, a remote coastal village in the Tumbes region of northern Peru. | 29.0 | 42† |
| Canto e Castro et al. | S | Portugal, using voluntary enrollment. | 43.3 | 34† |
| Kislaya et al. | L | Mainland Portugal and its autonomous regions, including Norte region, Centro region, Lisboa region, Alentejo region, Algarve region, Madeira, Azores. | 39.6* | 15 |

| | | | | |
|---|---|---|---|---|
| Talla et al. | S | All regions of Senegal. | 29.2 | 26 |
| Pérez-Gómez et al. | S | Spain, as part of a nationwide seroepidemiological investigation (ENE-COVID). | 48.4* | 17† |
| Richard et al. | S | The canton of Geneva, Switzerland, based on a representative sample of its residents. | 46.9 | 20 |
| Alsuwaidi et al. | L | The emirate of Abu Dhabi, including the regions of Abu Dhabi, Al Ain, and Al Dhafra. | 29.5* | 26† |
| Ward et al. | N | England, using a random sample from the National Health Service patient registry. | 52.5* | 22† |
| Ward et al. | N | England, using a random sample from the National Health Service patient registry. | 52.5* | 22† |
| Ward et al. | N | England, using a random sample from the National Health Service patient registry. | 52.2* | 22† |
| Sullivan et al. | N | The United States, using a national probability sample with at-home specimen collection. | 52.0* | 40 |
| Lamba et al. | N | California, USA, using a population-based sample to estimate statewide seroprevalence. | 51.0* | 41 |
| Chamberlain et al. | N | The state of Georgia, USA, using mailed specimen kits to estimate statewide seroprevalence. | 51.0* | 40 |
| Pathela et al. | S | New York City, USA. | 40.1* | 24 |

*Median age was calculated from the given age distribution.
†The recall period was not reported. We assumed it to be from 30 January 2020, when WHO declared COVID-19 a global health emergency, till the middle of the study duration.

**Table S2: Comparison of point estimates and median values of the proportion of asymptomatic individuals.** Point estimates of $\psi$ obtained using equation (1) and corresponding median values (and interquartile ranges) obtained by sampling the quantities in $Q$ (Methods). See Table 1 for study citations.

| Study | Point estimate | Median (IQR) |
|---|---|---|
| Menezes et al. | 0.29 | 0.27 (0.24, 0.29) |
| Silva et al. | 0.47 | 0.47 (0.45, 0.49) |
| Terças-Trettel et al. | 0.57 | 0.58 (0.56, 0.6) |
| Albuquerque et al. | 0.53 | 0.52 (0.49, 0.55) |
| Albuquerque et al. | 0.58 | 0.57 (0.54, 0.6) |
| Albuquerque et al. | 0.53 | 0.52 (0.49, 0.55) |
| Nwosu et al. | 0.87 | 0.87 (0.83, 0.9) |
| Vial et al. | 0.47 | 0.47 (0.44, 0.5) |
| Li et al. | 0.79 | 0.78 (0.76, 0.8) |
| Garay et al. | 0.64 | 0.64 (0.62, 0.66) |
| Serrano-Coll et al. | 0.84 | 0.84 (0.81, 0.87) |
| Espenhain et al | 0.53 | 0.53 (0.49, 0.56) |
| Carrat et al. | 0.34 | 0.41 (0.32, 0.47) |
| Rouquette et al. | 0.56 | 0.55 (0.47, 0.6) |
| Beaumont et al. | 0.31 | 0.31 (0.28, 0.34) |
| Santos-Hövener et al. | 0.44 | 0.47 (0.43, 0.51) |
| Weis et al. | 0.36 | 0.35 (0.29, 0.41) |
| Merkely et al. | 0.81 | 0.79 (0.71, 0.86) |
| Murhekar et al. | 0.98 | 0.98 (0.98, 0.98) |
| Selvaraju et al. | 0.96 | 0.96 (0.96, 0.97) |
| Kumar et al. | 0.94 | 0.94 (0.93, 0.94) |
| Sharma et al. | 0.99 | 0.99 (0.99, 0.99) |
| Sharma et al. | 1.00 | 0.997(0.995, 0.999) |
| Sharma et al. | 0.99 | 0.98 (0.98, 0.99) |
| Kumar et al. | 0.82 | 0.82 (0.79, 0.84) |
| Khan et al. | 0.96 | 0.96 (0.95, 0.96) |
| Poustchi et al. | 0.59 | 0.6 (0.57, 0.63) |
| Shadmani et al. | 0.73 | 0.73 (0.7, 0.75) |
| Heavey et al. | 0.37 | 0.36 (0.28, 0.42) |
| Melotti et al. | 0.31 | 0.3 (0.27, 0.33) |
| Pagani et al. | 0.32 | 0.35 (0.32, 0.38) |
| Ngere et al. | 0.89 | 0.89 (0.86, 0.92) |
| Abdul-Raheem et al. | 0.65 | 0.65 (0.61, 0.69) |
| Sagara et al. | 0.85 | 0.87 (0.84, 0.89) |
| Basto-Abreu et al. | 0.73 | 0.72 (0.71, 0.73) |
| Arnaldo et al. | 0.78 | 0.76 (0.68, 0.81) |
| Arnaldo et al. | 0.60 | 0.81 (0.67, 0.91) |
| Arnaldo et al. | 0.60 | 0.83 (0.72, 0.89) |
| Vos et al. | 0.18 | 0.17 (0.11, 0.22) |
| Okpala et al. | 0.59 | 0.58 (0.54, 0.62) |
| Nisar et al. | 0.99 | 0.98 (0.97, 0.99) |

| | | |
|---|---|---|
| Huamani et al. | 0.65 | 0.65 (0.62, 0.68) |
| Díaz-Vélez et al. | 0.77 | 0.76 (0.74, 0.79) |
| Reyes-Vega et al. | 0.59 | 0.6 (0.57, 0.62) |
| Moyano et al. | 0.71 | 0.71 (0.68, 0.74) |
| Canto e Castro et al. | 0.33 | 0.32 (0.28, 0.35) |
| Kislaya et al. | 0.51 | 0.6 (0.49, 0.68) |
| Talla et al. | 0.99 | 0.93 (0.88, 0.97) |
| Pérez-Gómez et al. | 0.26 | 0.42 (0.41, 0.43) |
| Richard et al. | 0.31 | 0.3 (0.28, 0.33) |
| Alsuwaidi et al. | 0.95 | 0.95 (0.94, 0.95) |
| Ward et al. | 0.26 | 0.33 (0.27, 0.36) |
| Ward et al. | 0.24 | 0.33 (0.26, 0.36) |
| Ward et al. | 0.18 | 0.32 (0.24, 0.36) |
| Sullivan et al. | 0.38 | 0.47 (0.4, 0.55) |
| Lamba et al. | 0.54 | 0.47 (0.34, 0.61) |
| Chamberlain et al. | 0.55 | 0.5 (0.38, 0.63) |
| Pathela et al. | 0.44 | 0.45 (0.44, 0.46) |

**Table S3. Distributions of sensitivity and specificity of assays used in serosurveys.** For each study in our analysis, the test assay name, type, manufacturer, the mean sensitivity ($\alpha$) and specificity ($\beta$), and their 95% confidence intervals ($\alpha_l, \alpha_u$) and ($\beta_l, \beta_u$), respectively, are listed. Parameters defining the corresponding distributions ($Beta(a, b)$ for sensitivity and $Beta(c, d)$ for specificity; Text S2) derived from the means and confidence intervals are also listed. See Table 1 for study citations.

| **Study** | **Test name** | **Test type**** | **Test manufacturer** | $\boldsymbol{\alpha}$ | $\boldsymbol{\alpha_l}$ | $\boldsymbol{\alpha_u}$ | $\boldsymbol{a}$ | $\boldsymbol{b}$ | $\boldsymbol{\beta}$ | $\boldsymbol{\beta_l}$ | $\boldsymbol{\beta_u}$ | $\boldsymbol{c}$ | $\boldsymbol{d}$ |
|---|---|---|---|---|---|---|---|---|---|---|---|---|---|
| Menezes et al. | SARS-CoV-2 rapid point-of-care test | LFIA | Wondfo Biotech Co., Guangzhou, China | 0.85 | 0.81 | 0.88 | 100 | 17.3 | 1.00 | 0.98 | 1.00 | 68.3 | 0.20 |
| Silva et al. | Elecsys® Anti-SARS-CoV-2 | CLIA | Roche Diagnostics | 0.89 | 0.79 | 0.96 | 47.9 | 6.1 | 1.00 | 0.99 | 1.00 | 100 | 0.10 |
| Terças-Trettel et al. | Liaison SARS-CoV-2 S1/S2 IgG | CLIA | DiaSorin | 0.97 | 0.96 | 0.99 | 100 | 1.8 | 0.99 | 0.98 | 0.99 | 100 | 0.60 |
| Albuquerque et al.* | SARS-CoV-2 Antibody test | LFIA | Wondfo Biotech Co., Guangzhou, China | 0.85 | 0.81 | 0.88 | 100 | 17.3 | 1.00 | 0.98 | 1.00 | 68.3 | 0.20 |
| Albuquerque et al.* | SARS-CoV-2 Antibody test | LFIA | Wondfo Biotech Co., Guangzhou, China | 0.85 | 0.81 | 0.88 | 100 | 17.3 | 1.00 | 0.98 | 1.00 | 68.3 | 0.20 |
| Albuquerque et al.* | SARS-CoV-2 Antibody test | LFIA | Wondfo Biotech Co., Guangzhou, China | 0.85 | 0.81 | 0.88 | 100 | 17.3 | 1.00 | 0.98 | 1.00 | 68.3 | 0.20 |
| Nwosu et al. | Panbio COVID-19 IgG rapid test | LFIA | Abbott Laboratories | 0.92 | 0.83 | 0.97 | 61.8 | 6.1 | 0.94 | 0.90 | 0.96 | 100 | 6.60 |
| Vial et al. | Elecsys® Anti-SARS-CoV-2 | ELISA | Roche Diagnostics | 0.99 | 0.97 | 1.00 | 100 | 0.8 | 1.00 | 1.00 | 1.00 | 100 | 0.10 |
| Li et al.† | Micro-neutralization (MN) assays | Neutralization assay | Author designed | 0.95 | 0.84 | 1.00 | 25.4 | 1.4 | 1.00 | 0.98 | 1.00 | 42.5 | 0.10 |
| Garay et al.‡ | INgezim® COVID 19 DR test | ELISA | Eurofins Ingenasa | 0.98 | 0.94 | 1.00 | 85.7 | 1.9 | 1.00 | 0.98 | 1.00 | 100 | 0.50 |
| Serrano-Coll et al.‡ | INgezim® COVID 19 DR test | ELISA | Eurofins Ingenasa | 0.98 | 0.94 | 1.00 | 85.7 | 1.9 | 0.99 | 0.97 | 1.00 | 100 | 0.70 |
| Espenhain et al | Wantai SARS-CoV-2 Total Ab ELISA | ELISA | Beijing Wantai Biological | 0.97 | 0.92 | 0.99 | 100 | 3.7 | 1.00 | 0.99 | 1.00 | 75.2 | 0.10 |
| Carrat et al. | Anti-SARS-CoV-2 ELISA IgG | ELISA | Euroimmun | 0.87 | 0.80 | 0.92 | 100 | 15.2 | 0.98 | 0.96 | 0.99 | 100 | 1.80 |
| Rouquette et al.γ | Anti-SARS-CoV-2 ELISA IgG | ELISA | Euroimmun | 1.00 | 0.92 | 1.00 | 11.6 | 0.1 | 0.98 | 0.92 | 1.00 | 61.5 | 1.80 |

| | | | | | | | | | | | | | |
|---|---|---|---|---|---|---|---|---|---|---|---|---|---|
| Beaumont et al. | Elecsys® Anti-SARS-CoV-2 | CLIA | Roche Diagnostics | 1.00 | 0.97 | 1.00 | 98.9 | 0.7 | 1.00 | 1.00 | 1.00 | 100 | 0.10 |
| Santos-Hövener et al.[¥] | Anti-SARS-CoV-2 ELISA IgG | ELISA | Euroimmun | 0.88 | 0.83 | 0.92 | 100 | 13.2 | 0.99 | 0.98 | 1.00 | 100 | 0.40 |
| Weis et al.[ε] | Multiple tests (See Text S2) | Multiple types | Multiple | 1.00 | 1.00 | 1.00 | 100 | 0.1 | 1.00 | 1.00 | 1.00 | 100 | 0.10 |
| Merkely et al.[€] | Abbott Architect SARS-CoV-2 IgG | CLIA | Abbott Laboratories | 1.00 | 0.95 | 1.00 | 19.5 | 0.1 | 1.00 | 0.99 | 1.00 | 97.7 | 0.10 |
| Murhekar et al.[€] | Abbott Architect SARS-CoV-2 IgG | CLIA | Abbott Laboratories | 1.00 | 0.95 | 1.00 | 19.5 | 0.1 | 1.00 | 0.99 | 1.00 | 97.7 | 0.10 |
| Selvaraju et al.[€] | Abbott Architect SARS-CoV-2 IgG | CLIA | Abbott Laboratories | 1.00 | 0.95 | 1.00 | 19.5 | 0.1 | 1.00 | 0.99 | 1.00 | 97.7 | 0.10 |
| Kumar et al.[€] | Abbott Architect SARS-CoV-2 IgG | CLIA | Abbott Laboratories | 1.00 | 0.95 | 1.00 | 19.5 | 0.1 | 1.00 | 0.99 | 1.00 | 97.7 | 0.10 |
| Sharma et al.[ε] | COVID Kawach IgG ELISA | ELISA | J. Mitra & Co. Pvt. Ltd | 0.92 | 0.77 | 0.99 | 21.7 | 2.1 | 0.98 | 0.88 | 1.00 | 29.6 | 1.00 |
| Sharma et al.[ε] | ErbaLisa COVID-19 IgG | ELISA | Erba Mannheim | 0.99 | 0.93 | 1.00 | 38.6 | 0.6 | 0.99 | 0.94 | 1.00 | 45.7 | 0.60 |
| Sharma et al.[ε] | ErbaLisa COVID-19 IgG | ELISA | Erba Mannheim | 0.99 | 0.93 | 1.00 | 38.6 | 0.6 | 0.99 | 0.94 | 1.00 | 45.7 | 0.60 |
| Kumar et al.[ε] | ICMR-NIV Anti-SARS CoV-2 Human IgG ELISA COVID Kavach – MERILISA | ELISA | Meril Diagnostics (Meril Life) | 0.93 | 0.79 | 1.00 | 22 | 1.8 | 1.00 | 0.99 | 1.00 | 99.3 | 0.10 |
| Khan et al. | Abbott Architect SARS-CoV-2 IgG | CLIA | Abbott Laboratories | 1.00 | 0.95 | 1.00 | 19.5 | 0.1 | 1.00 | 0.99 | 1.00 | 97.7 | 0.10 |
| Poustchi et al. | ELISA kit Pishtaz Teb Diagnostics | ELISA | Pishtaz Diagnostics Iran | 0.72 | 0.65 | 0.78 | 100 | 39.4 | 0.98 | 0.95 | 1.00 | 100 | 1.70 |
| Shadmani et al.[£] | ELISA kit Pishtaz Teb Diagnostics | ELISA | Pishtaz Diagnostics Iran | 0.94 | 0.80 | 0.98 | 24.3 | 1.9 | 0.98 | 0.94 | 1.00 | 84.4 | 1.90 |
| Heavey et al. | Abbott Architect SARS-CoV-2 IgG | CLIA | Abbott Laboratories | 0.94 | 0.86 | 0.98 | 61 | 4.3 | 1.00 | 0.99 | 1.00 | 100 | 0.10 |
| Melotti et al. | Abbott Architect SARS-CoV-2 IgG | CLIA | Abbott Laboratories | 1.00 | 0.95 | 1.00 | 19.9 | 0.1 | 1.00 | 0.90 | 1.00 | 14.7 | 0.20 |
| Pagani et al. | Prima Lab SA Covid-19 IgG Rapid test | LFIA | Prima Lab, Switzerland | 0.97 | 0.91 | 0.99 | 56.3 | 1.9 | 0.96 | 0.95 | 0.97 | 100 | 3.00 |

| | | | | | | | | | | | | | |
|---|---|---|---|---|---|---|---|---|---|---|---|---|---|
| Ngere et al. | Wantai SARS-CoV-2 Total Ab ELISA | ELISA | Beijing Wantai Biological | 0.94 | 0.92 | 0.99 | 100 | 4.4 | 1.00 | 0.92 | 1.00 | 12 | 0.10 |
| Abdul-Raheem et al.§ | VITROS anti-SARS- CoV-2 IgG antibody | CLIA | Ortho Clinical Diagnostics Inc. | 0.90 | 0.89 | 1.00 | 97.2 | 7.2 | 1.00 | 0.95 | 1.00 | 21.2 | 0.10 |
| Sagara et al. | Author designed (ELISA) -Spike | ELISA | — | 0.74 | 0.52 | 0.90 | 13.8 | 5.1 | 0.99 | 0.98 | 1.00 | 100 | 0.50 |
| Basto-Abreu et al. | Roche Elecsys Anti-SARS-CoV-2pan-immunoglobulin immunoassay test | CLIA | Roche Diagnostics | 0.92 | 0.89 | 0.95 | 100 | 8.1 | 1.00 | 0.97 | 1.00 | 100 | 0.60 |
| Arnaldo et al. | Qingdao Hightop Biotech IgM/IgG Duo | LFIA | Qingdao Hightop Biotech Co, Ltd, Shandong, China | 0.87 | 0.70 | 0.95 | 24.8 | 4.3 | 0.99 | 0.95 | 1.00 | 100 | 1.60 |
| Arnaldo et al. | Panbio COVID-19 IgG/ IgM Rapid Test Duo | LFIA | Abbott Laboratories | 0.83 | 0.66 | 0.93 | 24.5 | 5.4 | 0.98 | 0.94 | 0.99 | 100 | 2.30 |
| Arnaldo et al. | Panbio COVID-19 IgG/ IgM Rapid Test Duo | LFIA | Abbott Laboratories | 0.83 | 0.66 | 0.93 | 24.5 | 5.4 | 0.98 | 0.94 | 0.99 | 100 | 2.30 |
| Vos et al.¥ | — | ELISA | — | 0.84 | 0.76 | 0.90 | 81.3 | 15.6 | 1.00 | 0.99 | 1.00 | 97.7 | 0.10 |
| Okpala et al.¥£ | Realy Tech SARS-CoV-2 Antibodies Rapid Test Device | — | — | 0.98 | 0.95 | 0.99 | 100 | 2 | 0.99 | 0.93 | 1.00 | 39 | 0.70 |
| Nisar et al. | Elecsys® Anti-SARS-CoV-2 | CLIA | Roche Diagnostics | 0.97 | 0.95 | 0.98 | 100 | 2.1 | 1.00 | 0.99 | 1.00 | 100 | 0.10 |
| Huamani et al. | Elecsys® Anti-SARS-CoV-2 | CLIA | Roche Diagnostics | 0.97 | 0.95 | 0.98 | 100 | 2.1 | 1.00 | 0.99 | 1.00 | 100 | 0.10 |
| Díaz-Vélez et al. | Coretest COVID-19 IgM/IgG Ab Test | LFIA | Core Technology | 0.67 | 0.49 | 0.78 | 25.3 | 13.6 | 0.97 | 0.94 | 0.98 | 100 | 2.70 |
| Reyes-Vega et al.¥ | Standard Q COVID-19 IgM/IgG Duo rapid immunochromatography test kit | LFIA | SD Biosensor | 0.97 | 0.91 | 0.99 | 77.1 | 2.9 | 0.96 | 0.93 | 0.98 | 100 | 3.60 |
| Moyano et al. | Standard Q COVID-19 IgM/IgG Combo | LFIA | SD Biosensor | 0.99 | 0.95 | 1.00 | 67.8 | 1 | 0.99 | 0.96 | 1.00 | 100 | 1.30 |
| Canto e Castro et al.¥ | — | CLIA | Siemens Heathineers | 0.98 | 0.97 | 0.99 | 100 | 1.2 | 1.00 | 0.99 | 1.00 | 100 | 0.10 |
| Kislaya et al. | Anti-SARS-CoV-2 ELISA IgG,Wantai SARS-CoV-2 IgM ELISA | ELISA | Euroimmun,Beijing Wantai Biological | 0.96 | 0.91 | 0.98 | 98.4 | 4.7 | 0.99 | 0.97 | 0.99 | 100 | 0.80 |

| | | | | | | | | | | | | | |
|---|---|---|---|---|---|---|---|---|---|---|---|---|---|
| Talla et al.[ℇ] | Multiple types (See Text S2) | Multiple | Multiple | 0.81 | 0.62 | 0.93 | 19.5 | 5 | 0.99 | 0.92 | 1.00 | 42.7 | 0.90 |
| Pérez-Gómez et al. | Abbott Architect SARS-CoV-2 IgG | CMIA | Abbott Laboratories | 0.91 | 0.88 | 0.93 | 100 | 9.5 | 0.99 | 0.99 | 1.00 | 97.7 | 0.10 |
| Richard et al.[ℇ] | Anti-SARS-CoV-2 ELISA IgG | ELISA | Euroimmun | 0.93 | 0.79 | 1.00 | 22.1 | 1.9 | 1.00 | 0.99 | 1.00 | 99.3 | 0.10 |
| Alsuwaidi et al. | Liaison SARS-CoV-2 S1/S2 IgG,Elecsys® Anti-SARS-CoV-2 (N) | CLIA | DiaSorin,Roche Diagnostics | 0.97 | 0.96 | 0.99 | 100 | 1.8 | 0.99 | 0.98 | 0.99 | 100 | 0.60 |
| Ward et al. | Coronavirus Antibody Rapid Test | LFIA | Fortress Diagnostics | 0.84 | 0.71 | 0.94 | 31.4 | 6.1 | 0.99 | 0.97 | 0.99 | 100 | 0.80 |
| Sullivan et al. | Platelia SARS-CoV-2 Total Ab assay | ELISA | Bio-rad | 0.98 | 0.90 | 1.00 | 33.3 | 1 | 0.99 | 0.98 | 1.00 | 100 | 0.80 |
| Lamba et al.[ℇ] | Platelia SARS-CoV-2 Total Ab assay | ELISA | Bio-rad | 0.92 | 0.77 | 0.99 | 21.2 | 2 | 1.00 | 0.96 | 1.00 | 100 | 0.80 |
| Chamberlain et al.[ℇ] | Platelia SARS-CoV-2 Total Ab assay | ELISA | Bio-rad | 0.92 | 0.77 | 0.99 | 21.2 | 2 | 1.00 | 0.96 | 1.00 | 100 | 0.30 |
| Pathela et al. | Liaison SARS-CoV-2 S1/S2 IgG | CLIA | DiaSorin | 0.98 | 0.96 | 0.99 | 100 | 1.7 | 0.99 | 0.98 | 0.99 | 58.9 | 0.50 |

*Parameters obtained from Menezes et al., who used the same assay.

†Seropositivity in Li et al. was defined by a micro-neutralization (MN) titer ≥4; Li et al. validated the MN assay on healthy and non-reactive sera and observed no detectable neutralizing antibodies in controls (supporting very high specificity). The 95% CI was calculated using Wilson score interval. The study did not report an empirical MN sensitivity. Based on (a) the Hong Kong MN results cited by Li et al. (8/9 detected using a higher MN threshold) and (b) Li et al.'s use of a lower threshold (≥4) to reduce false negatives, we assume an MN sensitivity of 0.95.

‡Mean of $\alpha$ and $\beta$ obtained from Mattar et al., study cited by Garay et al.; corresponding CIs computed using the Wilson score interval.

[γ]Multiple sources cited for $\alpha$ and $\beta$; estimates from Beavis et al.[14] were used owing to their larger sample size.

¥95% CI for $\alpha$ and $\beta$ calculated using Wilson score interval.

[ℇ]Mean and 95% CI of $\alpha$ and $\beta$ were calculated as described in Text S2.

€95% CIs obtained from manufacturer data (Abbott SARS-CoV-2 assay): https://www.corelaboratory.abbott/us/en/offerings/segments/infectious-disease/sars-cov-2.html#isi

[£]CIs for $\alpha$ and $\beta$ calculated using Wilson score interval and manufacturer validation data given in Poustchi et al.

[§]CIs for α and β obtained from FDA document: https://www.accessdata.fda.gov/cdrh_docs/presentations/maf/maf3371-a001.pdf

**LFIA - Lateral flow immunoassay, CLIA - Chemiluminescent immunoassay, ELISA - Enzyme-linked immunosorbent Assay, CMIA - Chemiluminescent microparticle immunoassay

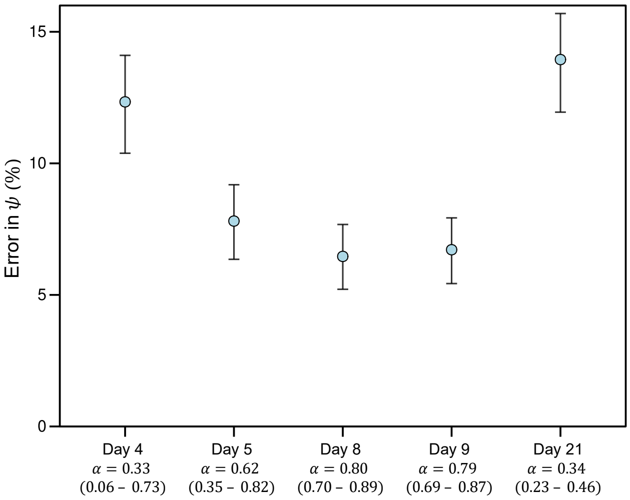


**Fig. S1. Effect of temporal variation in test characteristics on the accuracy of $\psi$.** Synthetic data were generated (Text S1) to evaluate how changing $\alpha$ over the course of infection influences the accuracy of our estimate of $\psi$. The percentage error in $\psi$, defined as $(\psi - \psi_{\text{true}})/\psi_{\text{true}} \times 100$, where $\psi_{true}$ is the true asymptomatic proportion used to generate the synthetic data, is estimated as a function of time from exposure (typically 4 days before symptom onset). The values of $\alpha$ at the different time points, obtained from Kucirka et al.[1], are also shown. See Text S1 for details.

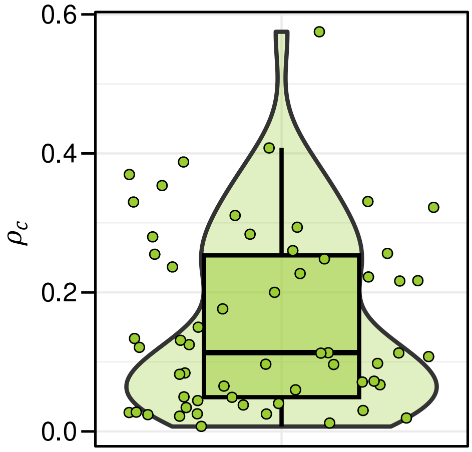


**Fig. S2. Distribution of crude seroprevalence across serosurveys.** The distribution of the estimates (symbols) of $\rho_c$ extracted from 50 serosurveys (Table 1). The width of the violin is proportional to the density of estimates of $\rho_c$. The box plot depicts the quartiles, the central horizontal line denotes the median, whiskers represent 1.5 times the interquartile range beyond the box.

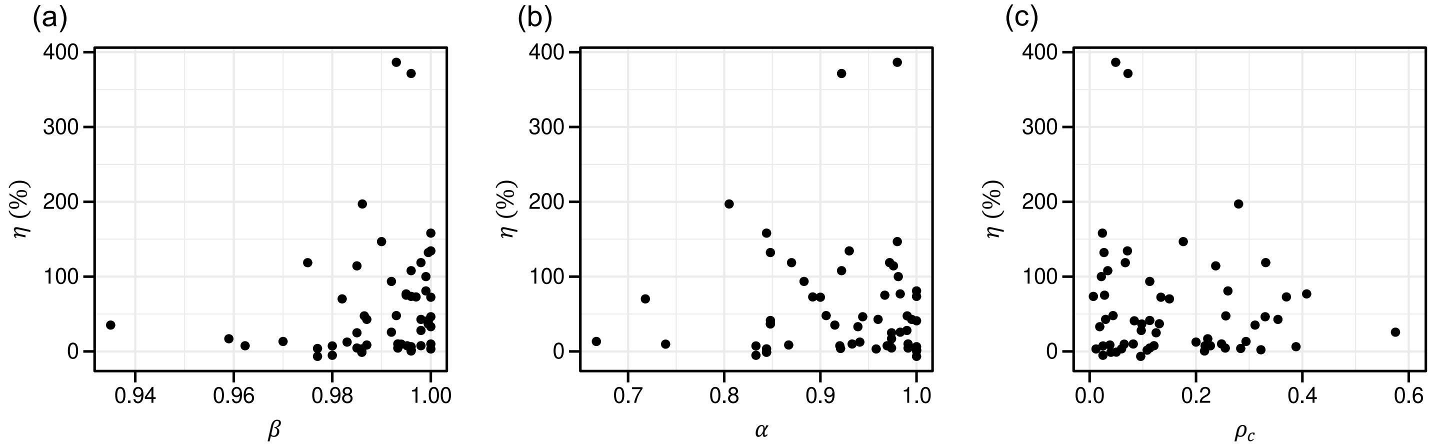


**Fig. S3. Correlations between $\eta$ and quantities in $Q$**. Pairwise correlation between (a) $\eta$ and $\beta$, (b) $\eta$ and $\alpha$, and (c) $\eta$ and $\rho_c$, for the serosurveys we studied (Table 1). $r_s$ denotes Spearman's correlation coefficient. The observed correlations were: (a) $r_s$ = 0.28, P = 0.03, (b) $r_s$ = −0.06*,* P = 0.63; and (c) $r_s$ = -0.02, P = 0.87. (See Fig. 4 for factors associated with $\eta$.)

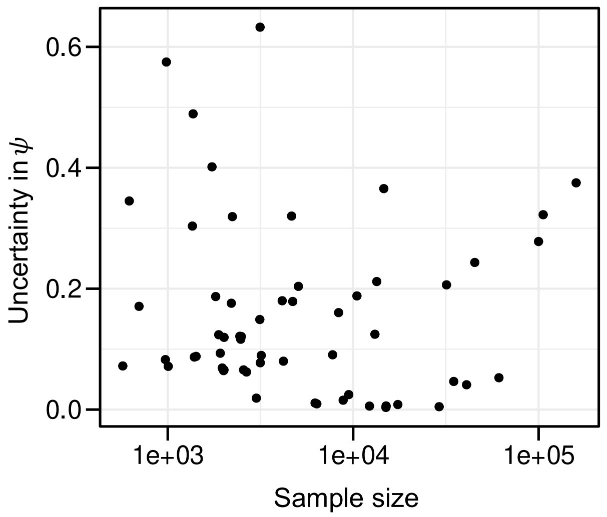


**Fig. S4. Influence of sample size on estimates of $\boldsymbol{\psi}$.** The uncertainty in the estimates of $\psi$, estimated as the interquartile range of $\psi$ normalized by the median, $(Q_3 - Q_1)/\text{median}(\psi)$, versus serosurvey sample size for the 58 estimates we examined (Table 1). The Spearman correlation coefficient, $r_s$=-0.17 (P=0.2).

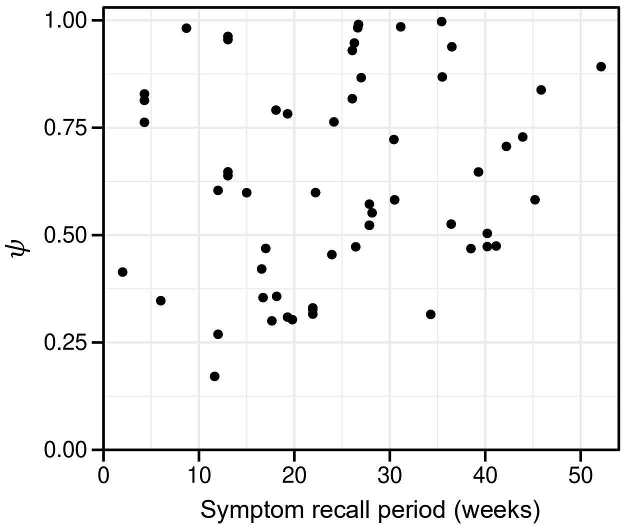


**Fig. S5. Influence of symptom recall period on estimates of $\psi$.** The estimate of $\psi$ obtained using our formalism plotted against the symptom recall period used in the serosurvey, shown for all the serosurveys we examined (Table 1). Spearman's correlation coefficient, $r_s$=0.18 (P=0.17).

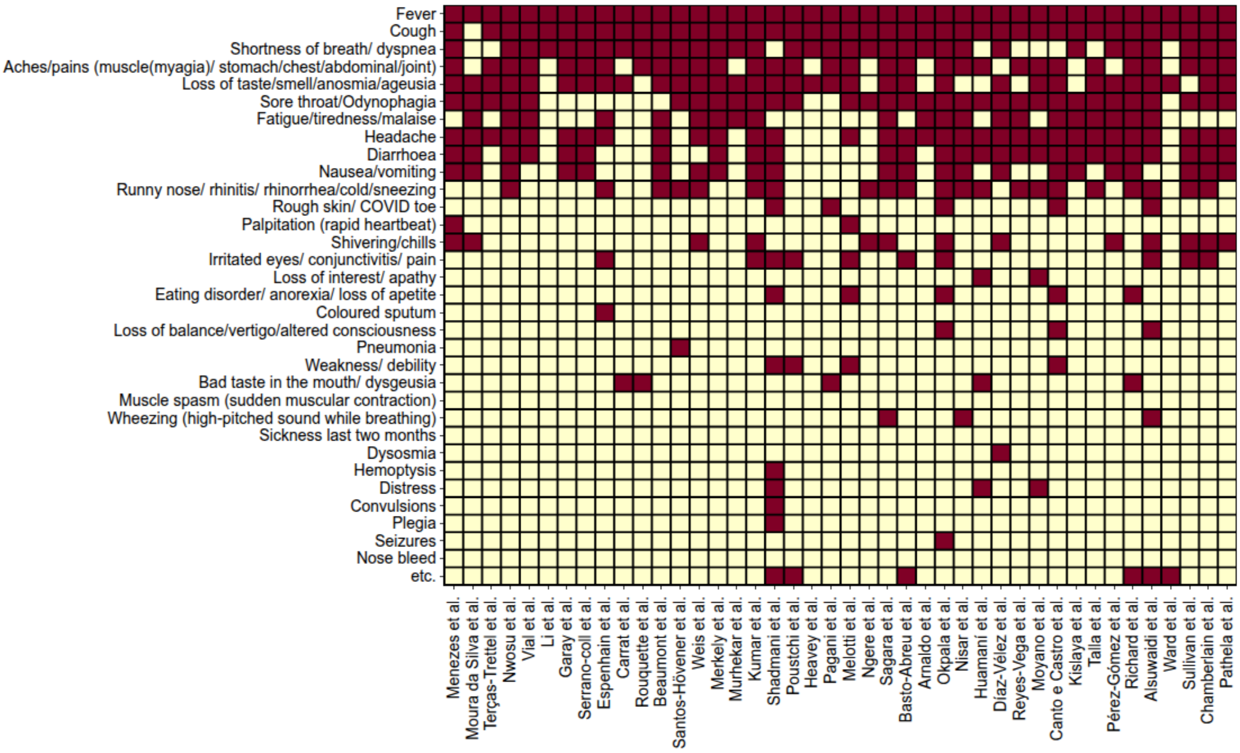


**Fig. S6. Symptom sets.** Symptom sets used by serosurveys in our analysis. Details of the surveys are in Table 1. Symptoms included in the respective surveys are colored red and those excluded are yellow. The surveys reported by Albuquerque et al., Selvaraju et al., Kumar et al. (Chennai study), Sharma et al., Khan et al., Abdul-Raheem et al., Lamba et al. did not report the symptoms considered (see Table 1 for citations).

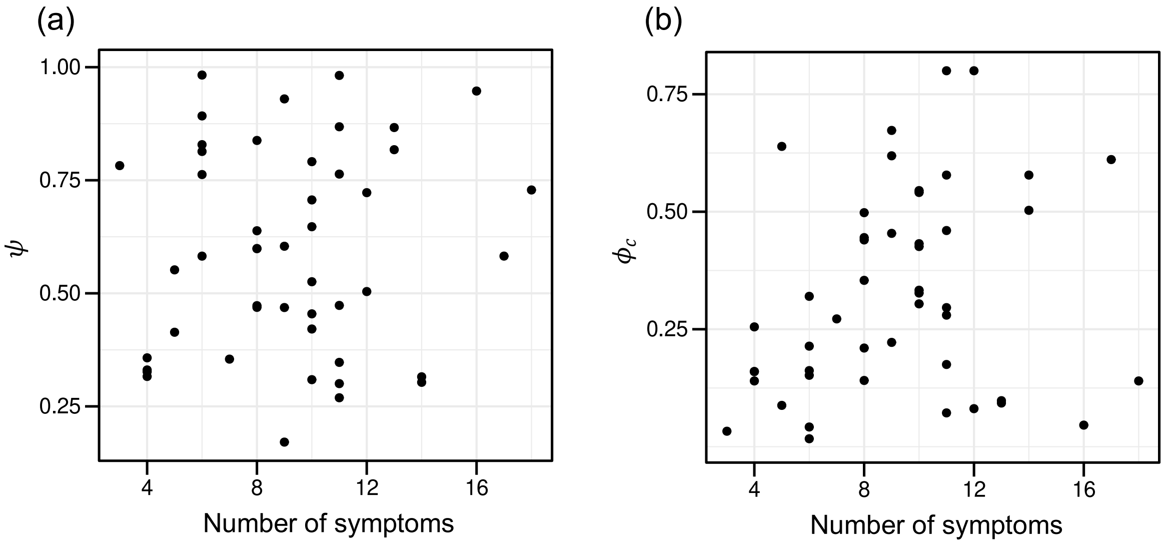


**Fig. S7. Robustness to heterogeneity in symptom sets.** Estimates of (a) $\psi$ and (b) $\phi_c$ versus the number of symptoms used in individual studies (see Fig. S6). No significant association was found in either case. (a) Spearman's correlation coefficient, $r_s$=0.06 (P=0.70). (b) $r_s$=0.25 (P=0.09).

**Supplementary references**


1. Kucirka, L., Lauer, S., Laeyendecker, O., Boon, D. & Lessler, J. Variation in false-negative rate of reverse transcriptase polymerase chain reaction–based SARS-CoV-2 tests by time since exposure. *Ann. Intern. Med.* **173**, 262-267 (2020).
2. Mair, M.D., *et al.* A systematic review and meta-analysis comparing the diagnostic accuracy of initial RT-PCR and CT scan in suspected COVID-19 patients. *Br. J. Radiol.* **94**, 20201039 (2021).
3. Dan, J.M., *et al.* Immunological memory to SARS-CoV-2 assessed for up to 8 months after infection. *Science* **371**, eabf4063 (2021).
4. Okpala, O.V., *et al.* Population seroprevalence of SARS-CoV-2 antibodies in Anambra State, South-East, Nigeria. *Int. J. Infect. Dis.* **110**, 171-178 (2021).
5. Li, Z., *et al.* Antibody seroprevalence in the epicenter Wuhan, Hubei, and six selected provinces after containment of the first epidemic wave of COVID-19 in China. *Lancet Reg. Health. West. Pac.* **8**, 100094 (2021).
6. Sharma, N., *et al.* The seroprevalence of severe acute respiratory syndrome coronavirus 2 in Delhi, India: a repeated population-based seroepidemiological study. *Trans. R. Soc. Trop. Med. Hyg.* **116**, 242-251 (2022).
7. Kumar, D., *et al.* Seroprevalence of anti SARS-CoV-2 IgG antibodies among adults in Jammu district, India: A community-based study. *Indian J. Med. Res.* **155**, 171-177 (2022).
8. Richard, A., *et al.* Seroprevalence of anti-SARS-CoV-2 IgG antibodies, risk factors for infection and associated symptoms in Geneva, Switzerland: a population-based study. *Scand. J. Public Health* **50**, 124-135 (2022).
9. Lamba, K., *et al.* SARS-CoV-2 cumulative incidence and period seroprevalence: Results from a statewide population-based serosurvey in California. *Open Forum Infect. Dis.* **8**, ofab379 (2021).
10. Chamberlain, A.T., *et al.* Cumulative incidence of SARS-CoV-2 infections among adults in Georgia, United States, August to December 2020. *J. Infect. Dis.* **225**, 396-403 (2022).
11. Gelman, A., *et al. Bayesian Data Analysis*, (Chapman and Hall/CRC, New York, 2013).
12. Weis, S., *et al.* Antibody response using six different serological assays in a completely PCR-tested community after a coronavirus disease 2019 outbreak-the CoNAN study. *Clin. Microbiol. Infect.* **27**, 470 e471-470 e479 (2021).
13. Talla, C., *et al.* Seroprevalence of anti-SARS-CoV-2 antibodies in Senegal: a national population-based cross-sectional survey, between October and November 2020. *IJID Reg* **3**, 117-125 (2022).
14. Beavis, K.G., *et al.* Evaluation of the EUROIMMUN Anti-SARS-CoV-2 ELISA Assay for detection of IgA and IgG antibodies. *J. Clin. Virol.* **129**, 104468 (2020).
